# Testing the Triple Network Model of Psychopathology in a Transdiagnostic Neurodevelopmental Cohort

**DOI:** 10.1101/2022.05.05.22274709

**Authors:** Jonathan S. Jones, The CALM Team, Amelia Leyland-Craggs, Duncan E. Astle

## Abstract

**Background:** The triple network model of psychopathology posits that altered connectivity between the Salience (SN), Central Executive (CEN), and Default Mode Networks (DMN) may underlie neurodevelopmental conditions. However, this has yet to be tested in a transdiagnostic sample of youth.

**Methods:** We investigated triple network connectivity in a sample of 175 children (60 girls) that represent a heterogeneous population who are experiencing neurodevelopmental difficulties in cognition and behavior, and 60 comparison children (33 girls) without such difficulties. Hyperactivity/impulsivity and inattention were assessed by parent-report and resting-state functional Magnetic Resonance Imaging data were acquired. Functional connectivity was calculated between independent network components and regions of interest. We then examined whether connectivity between the SN, CEN and DMN was dimensionally related to hyperactivity/impulsivity and inattention, whilst controlling for age, gender, and motion.

**Results:** Hyperactivity/impulsivity was associated with decreased segregation between the SN, CEN, and DMN in at-risk children, whereas it was associated with increased segregation of the CEN and DMN in comparison children. We replicated these effects in networks and regions derived from an adult parcellation of brain function and when using increasingly stringent exclusion criteria for in-scanner motion.

**Conclusions:** Triple network connectivity characterizes transdiagnostic neurodevelopmental difficulties with hyperactivity/impulsivity. This may arise from delayed network segregation, difficulties sustaining CEN activity to regulate behavior, and/or a heightened developmental mismatch between neural systems implicated in cognitive control relative to those implicated in reward/affect processing.

---

Neurodevelopmental conditions affect up to 10% of children (1) but many more require additional support at school (2). These conditions vary widely in scope and impact, often being characterized in terms of cognition, behavior, communication, mental health, academic attainment, and lived experience. In recent years there has been a gradual shift away from seeking single common causes of individual disorder categories, towards the identification of mechanisms that may explain neurodevelopmental characteristics which span diagnostic boundaries (3–6). One prominent example is the ‘triple network model of psychopathology’ (hereafter termed the ‘triple network model’), which postulates that atypical functional interactions between the Salience (SN), Central Executive (CEN) and Default-Mode Networks (DMN) underlie a range of neurodevelopmental, mental health, psychiatric, and neurodegenerative characteristics (7). This transdiagnostic model suggests that some common difficulties in mood and cognition could reflect alterations in the coupling between these networks. Support for this theory has largely come from independent diagnostically-based case-control studies, showing group differences in network interactions or correlations between these interactions and symptom severity. However, the model has yet to be tested in a transdiagnostic sample of children. This is an important next step because, if correct, the model would predict that neurodevelopmental difficulties reflect differential interactions between these three networks. Here, we test the triple network model in a broad heterogeneous sample of children with a range of diagnostic statuses who were referred for cognitive and behavioral difficulties. Specifically we tested whether functional connectivity between the SN, CEN and DMN is significantly associated with inattention and hyperactivity/impulsivity.

Interactions between the SN, CEN and DMN may help to characterize neurodevelopmental diversity because of their critical roles in higher-order cognition and goal-directed behavior (7). They are emergent properties of adult brain function (8,9), develop in childhood (10), and are linked to deviations in cognitive development (11). The CEN is a lateral fronto-parietal network primarily consisting of the lateral prefrontal (lPFC) and posterior parietal cortices, and is broadly involved in cognitive control (12). The DMN primarily comprises of the medial prefrontal cortex (mPFC), posterior cingulate (PC), precuneus and angular gyrus, and it is involved in emotion processing, self-referential thought, social cognition, and episodic memory (13). Finally, the SN primarily comprises of the dorsal anterior cingulate (AC) and anterior insula (AI), and is involved in the detection of salient internal and external stimuli to adaptively guide behavior (14). The CEN is activated during externally-oriented cognitively demanding tasks whereas the DMN is typically deactivated. The SN has been shown to mediate this switch between cognition of external and internal stimuli (15).

To date, the triple network model has only been examined in case-control studies of Attention Deficit Hyperactivity Disorder (ADHD) and autism. These studies have reported functional connectivity differences within and between the SN, CEN, and DMN, which scale with symptom severity (16–21). However, this altered functional connectivity may reflect a broader phenotype of behavioral characteristics that commonly occur in both ADHD and autism, rather than a diagnostic status per se. For example, hyperactivity/impulsivity and inattention are common across neurodevelopmental conditions (22–25), occurring in as many as 76% of autistic children (26) and those without any formal diagnosis (27). Transdiagnostic studies, with more inclusive sampling frameworks, therefore allow researchers to associate particular aspects of neurobiology with behavioral characteristics.

In the current study, we used a large transdiagnostic sample of children who experience difficulties in cognition and behavior, with variable scope and impact (27–34). Children were referred to the sample by professionals in children’s educational or clinical services, and are at elevated risk of educational underachievement (35), underemployment (36), and mental health difficulties (37). Hereafter, we refer to these children as neurodevelopmentally ‘at risk’. We also included a comparison sample of children not referred on the basis of neurodevelopmental difficulties. We then tested whether functional connectivity between the SN, CEN and DMN is associated with parent-reported hyperactivity/impulsivity and inattention in each sample.

## Methods and Materials

### Sample Characteristics

Behavioral and resting-state functional Magnetic Resonance Imaging (fMRI) data were available for 343 children and adolescents who opted to take part in the MRI portion of the Centre for Attention, Learning and Memory project (31). Neurodevelopmentally at-risk children were identified by education and clinical practitioners as struggling in the areas of attention, learning, language, and memory. Comparison children were recruited from the same schools without such difficulties. Children with high motion scans (*n=*108) were excluded from the analysis (see ‘fMRI Preprocessing’ for details). The final sample consisted of 235 children and adolescents aged 5-17 years: 175 at-risk and 60 comparison children (see Table 1).

**Table 1.** Group Characteristics.

|  | All (*n*=235) | At-risk (*n*= 175) | Comparison (*n*=60) |
| --- | --- | --- | --- |
| Age in years: *M* (*SD*) | 10.79 (2.17) | 10.72 (2.20) | 11.02 (2.08) |
| Boys: *n* | 142 (60.42%) | 115 (65.71%) | 27 (45%) |
| Girls: *n* | 93 (39.57%) | 60 (34.29%) | 33 (55%) |
| Ethnicity: *n* |  |  |  |
| Asian/Asian British | 0 (0%) | 0 (0%) | 0 (0%) |
| Black/African/Caribbean/Black British | 1 (0.88%) | 0 (0%) | 1 (2.7%) |
| Mixed/Multiple Ethnic Groups | 10 (8.85%) | 8 (10.53%) | 2 (5.41%) |
| White | 102 (90.27%) | 68 (89.47%) | 34 (91.89%) |
| Any diagnosis<sup>a</sup>: *n* | 69 (29.36%) | 66 (37.71%) | 3 (5%) |
| ADHD: *n* | 33 (14.04%) | 32 (18.29%) | 1 (1.67%) |
| Autism: *n* | 13 (5.53%) | 13 (7.42%) | 0 (0%) |
| Dyslexia: *n* | 19 (8.09%) | 17 (9.71%) | 2 (3.33%) |
| IMD: *M* (*SD*) | 21505 (7771) | 20421 (7713) | 24556 (7152) |
| Hyperactivity/Impulsivity: *M* (*SD*) | 8.12 (5.83) | 9.65 (5.59) | 3.65 (3.89) |
| Inattention: *M* (*SD*) | 9.49 (5.08) | 11.47 (3.68) | 3.7 (4.08) |
| SN-CEN FC: *M* (*SD*) | 0.10 (0.18) | 0.11 (0.18) | 0.05 (0.17) |
| SN-DMN FC: *M* (*SD*) | 0.16 (0.23) | 0.15 (0.23) | 0.17 (0.22) |
| CEN-DMN FC: *M* (*SD*) | 0.17 (0.18) | 0.18 (0.18) | 0.16 (0.17) |
| In-scanner motion: *M* (*SD*) | 0.20 (0.09) | 0.21 (0.09) | 0.16 (0.08) |

*Note.* Age at brain scan. The Index of Multiple Deprivation (IMD) was available for 229 children. Ethnicity data were available for 113 children. <sup>a</sup>Any neurodevelopmental or mental health diagnosis. Attention Deficit Hyperactivity Disorder (ADHD), Salience Network (SN), Central Executive Network (CEN), Default Mode Network (DMN), and Functional Connectivity (FC).

### Measures

#### Hyperactivity/Impulsivity and Inattention

The Conners Parent Rating Short Form 3^rd^ Edition is a validated and reliable parent questionnaire of behavior in childhood that is widely used in clinical contexts (38). Parents or carers rated the frequency of 11 behavioral items over the past month that corresponded to the two scales of hyperactivity/impulsivity and inattention. The raw total score of each scale was used in subsequent analyses.

### Image Acquisition

Magnetic resonance imaging data were acquired at the MRC Cognition and Brain Sciences Unit, University of Cambridge. All scans were obtained on a Siemens 3T Prisma-fit system (Siemens Healthcare, Erlangen, Germany), using a 32-channel quadrature head coil.

In the resting-state fMRI, 270 T2*-weighted whole-brain echo planar images (EPIs) were acquired over nine minutes (time repetition [TR] = 2s; time echo [TE] = 30ms; flip angle = 78 degrees, 3×3×3mm). The first 4 volumes were discarded to ensure steady state magnetization. Participants were instructed to lie still with their eyes closed and to not fall asleep. For registration of functional images, T1-weighted volume scans were acquired using a whole-brain coverage 3D Magnetization Prepared Rapid Acquisition Gradient Echo (MP-RAGE) sequence acquired using 1-mm isometric image resolution (TR = 2.25s, TE = 2.98ms, flip angle = 9 degrees, 1×1×1mm).

### fMRI Pre-processing

The data were minimally pre-processed in fMRIPrep version 1.5.0 (39), which implements slice-timing correction, rigid-body realignment, boundary-based co-registration to the structural T1, segmentation, and normalization to the MNI template. The data were then smoothed by 6mm full-width at half-maximum. Strategies to denoise motion and physiological artefacts were evaluated in fmridenoise (40). The most effective strategy included regression of 24 head motion parameters (six rigid body realignment parameters, their squares, their derivatives, and their squared derivatives), 10 aCompCor components derived from the WM and CSF signal (41), linear and quadratic trends, motion spikes (framewise displacement >0.5mm; 42), and a band-pass filter between 0.01-0.1Hz (see 27, for details). Simultaneous confound regression was performed in the Nipype (version 1.2.0) implementation of AFNI’s 3dTproject (43). Children who moved substantially during the scan were excluded from the analysis: first on the basis of high average motion (mean framewise displacement >0.5mm, *n*=89) and then on the number of motion spikes (>20% spikes, *n*=19), where few temporal degrees of freedom would have remained. The final sample included 235 children. Average in-scanner motion was 0.2mm (*SD*=0.09mm).

### Resting-state Networks

Spatial components of the denoised resting-state data were identified using canonical Independent Components Analysis on the whole sample using nilearn 0.7.0, with the number of components set to 30 (44). We visually identified five components that corresponded with the SN, CEN, and DMN. The SN component included: the bilateral AC, medial orbitofrontal cortex (mOFC), bilateral ventral AI, caudate head, nucleus accumbens, and globus pallidus. The left CEN component included: the left lPFC, right lPFC, left frontal pole, left medial superior frontal gyrus, and left intraparietal sulcus (IPS). The right CEN component included: the right lPFC, right frontal pole, right medial superior frontal gyrus, and right IPS / inferior parietal lobe. The DMN component included: the mPFC, retrosplenial cortex, bilateral angular gyrus, bilateral superior frontal cortex and hippocampus. Finally, the posterior DMN component included: the bilateral PC / precuneus and bilateral IPS. Binary masks were created for each component and the DMN masks were combined (see Figure 1). Time series were extracted from the four network masks and correlated using Pearson correlations to give a measure of functional connectivity between each pair of networks. Functional connectivity of the CEN with the SN and DMN was estimated as the average of the right and left CEN, respectively.

**Figure 1.**
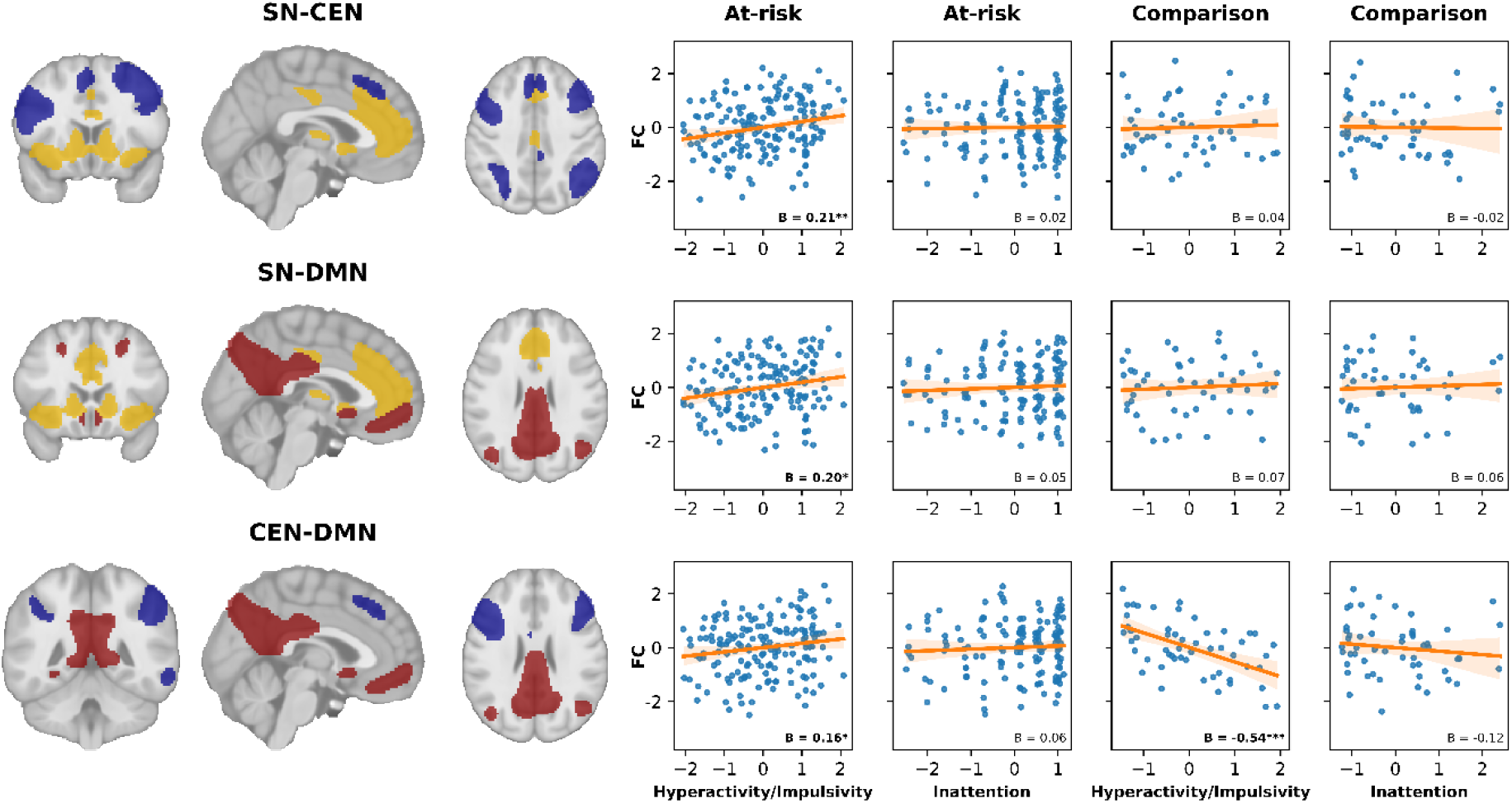
Associations between triple network connectivity and neurodevelopmental difficulties. Network masks derived from the ICA (left) are shown for the SN (yellow), CEN (blue), and DMN (red). Within-group partial regression plots (right) display the dimensional relations between behavioral difficulties and functional connectivity after age, age^2^, gender, and in-scanner motion have been regressed out. The shaded area around the regression line shows 95% confidence intervals from 1000 bootstrapped resamples. Salience Network (SN), Central Executive Network (CEN), and Default Mode Network (DMN). \*\*\**p<*0.001, \*\**p<*0.01, \**p<*0.05

### Adult Networks and Regions of Interest

We also investigated functional connectivity between 14 key canonical regions of the SN, CEN, and DMN derived from a 100 region 17 networks parcellation of adult resting-brain function (45). This included bilateral regions of the SN (dorsal AI and dorsal AC), CEN (lPFC and IPS), and DMN (mPFC and PC / precuneus). In our analyses of the SN, we also included a region of the ventral AI; although this was assigned to the adult DMN, it aligns with both the data-driven SN component and the original description of the SN (46,47). We analyzed inter-regional functional connectivity within every network and between every pair of networks. Within each set of analyses, we corrected for multiple comparisons using the False Discovery Rate (FDR) Benjamini-Hochberg procedure (48). Finally, we analyzed functional connectivity between the adult networks by combining the regions into four network masks and correlating the extracted time-series: the dorsal SN (dorsal AI and dorsal AC), ventral SN (ventral AI and dorsal AC), CEN, and DMN.

### Statistical Analyses

Dimensional relations between connectivity and behavior were investigated within each sample using ordinary least squares regression whilst controlling for age, age^2^, gender, and in-scanner motion. We then examined whether these effects were evident at the group level in neurodevelopmentally at-risk children and those diagnosed with combined type ADHD, relative to the comparison sample. Finally, we investigated whether the dimensional associations between connectivity and behavior differed between neurodevelopmentally at-risk and comparison children. Univariate statistical outliers greater than or less than three times the median absolute deviation were excluded from each analysis in a casewise manner, as in previous studies (19). All analysis code is available at: https://osf.io/7mptz/

## Results

### Triple Network Connectivity

Children with the most marked hyperactivity/impulsivity had decreased functional segregation between the SN, CEN, and DMN (see Figure 1 and Tables S1-S2). Within the neurodevelopmentally at-risk sample, hyperactivity/impulsivity was significantly associated with greater functional connectivity between the SN-CEN (*n=*172, *β=*0.21, *p=*0.006), SN-DMN (*n*=164, β=0.20, p=0.013), and CEN-DMN (*n=*166, *β=*0.16, *p=*0.042). However, these associations were not apparent in the comparison sample; instead, hyperactivity/impulsivity was significantly associated with reduced functional connectivity between the CEN-DMN (*n=*53, *β=*-0.54, *p=*6.7E-05), and this brain-behavior association significantly differed between the groups (*n=*219, *β=*0.65, *p*=4.89E-05).

We then evaluated whether these effects generalized to a common parcellation of adult resting brain function (see Tables S3-S4). In the at-risk sample, hyperactivity/impulsivity was again significantly associated with greater CEN-DMN connectivity (*n=*169, *β=*0.21, *p=*0.009) and greater SN-DMN connectivity (ventral SN: *n=*171, *β=*0.2, *p=*0.009; dorsal SN: *n=*166, *β=*0.15, *p=*0.05), but not SN-CEN connectivity (ventral SN: *n=*160, *β=*0.03, *p=*0.723; dorsal SN: *n=*164, *β=*-0.07, *p=*0.373). As before, there was a significant group interaction on the association between hyperactivity/impulsivity and CEN-DMN connectivity (*n=*222, *β=*0.37, *p=*0.021).

### Group Differences

Group differences also implicated elevated SN-CEN functional connectivity with neurodevelopmental difficulties (see Tables S5-S6). Specifically, SN-CEN connectivity was greater in children diagnosed with combined type ADHD (*n=*32, *M* = 0.16, *SD* = 0.13) than comparison children (*n=*58, *M* = 0.06, *SD* = 0.16); *β=*0.56, *p=*0.023. SN-CEN connectivity was also greater in neurodevelopmentally at-risk children (*n=*172, *M* = 0.11, *SD* = 0.17) than comparison children (*n=*59, *M* = 0.06, *SD* = 0.16); however, this was not significant (*β=*0.19, *p=*0.220).

### Regional Connectivity

In line with the network level effects, decreased functional segregation between core regions of the SN, CEN, and DMN was associated with more marked hyperactivity/impulsivity in the at-risk sample, but not the comparison sample (see Figure 2 and Tables S7-12). Between the CEN and DMN, hyperactivity/impulsivity was significantly associated with greater connectivity between the right lPFC and right mPFC (*n=*172, *β=*0.25, *p=*0.008 FDR-corrected) and the right lPFC and left mPFC (*n=*172, *β=*0.29, *p=*0.003 FDR-corrected). Between the SN and DMN, hyperactivity/impulsivity was associated with greater connectivity between the right dorsal AC and left PC (*n=*170, *β=*0.26, *p=*0.028 FDR-corrected).

**Figure 2.**
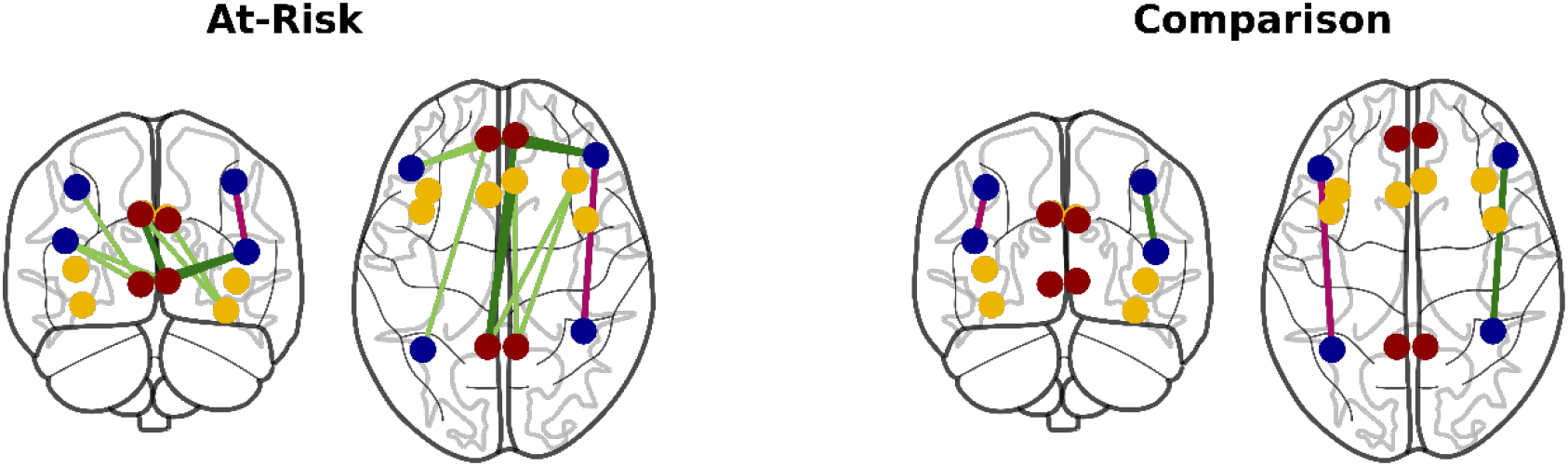
Associations between hyperactivity/impulsivity and inter-regional functional connectivity in the Salience (yellow), Central Executive (blue), and Default Mode Networks (red). Green lines indicate positive associations and pink lines indicate negative associations. Borderline significant associations are shown in lighter colors with less weight (*p<*0.073).

Within networks, decreased functional integration within the CEN and increased integration within the DMN was associated with more marked hyperactivity/impulsivity in the at-risk sample (see Tables S13-18). In the CEN, hyperactivity/impulsivity was associated with decreased connectivity between the right lPFC and right IPS (*n=*162, *β=*-0.22, *p=*0.04 FDR-corrected). In the DMN, hyperactivity/impulsivity was associated with increased connectivity between the right mPFC and left PC (*n=*163, *β=*0.22, *p=*0.042 FDR-corrected). In the comparison sample, the associations between regional CEN integration and hyperactivity/impulsivity were mixed. Consistent with the at-risk sample, greater connectivity between the *left* lPFC and *left* IPS was associated with *less* hyperactivity/impulsivity (*n=*53, *β=*-0.35, *p=*0.046 FDR-corrected). But in direct contrast with the at-risk sample, and as indicated by significant group interaction (*n=*216, *β=*0.44, *p=*0.045 FDR-corrected), greater connectivity between the *right* lPFC and *right* IPS was associated with *greater* hyperactivity/impulsivity in the comparison sample (*n=*54, *β=*0.36, *p=*0.046 FDR-corrected).

### Robustness Analyses

As our findings pertained to hyperactivity, we opted to further rule out the confounding effects of in-scanner motion by repeating our main analyses after excluding children with mean framewise displacement above 0.15-0.45mm. Overall, effect sizes were similar and, despite limited power in these smaller samples, we were able to replicate our findings at multiple thresholds (see Tables S19-26).

### Exploring the group*behavior interaction in CEN-DMN connectivity

We conducted post-hoc tests to explore the group interaction of hyperactivity/impulsivity on CEN-DMN connectivity (see Supplementary Materials). We first investigated the hypothesis that the functional segregation of the CEN and DMN may diverge with age in the at-risk sample relative to the comparison sample. We found that CEN-DMN connectivity seemed to linearly increase with age in the comparison sample (*n=*52, *β=*0.3, *p=*0.057), whereas it quadratically decreased in younger at-risk children before flattening in older children (*n=*169, *β=*0.16, *p=*0.051; see Figure S1). Second, we explored whether greater CEN-DMN connectivity may be cognitively advantageous in the comparison sample. Indeed, fluid intelligence was significantly positively associated with CEN-DMN connectivity (*n=*53, *β=*0.27, *p=*0.051) and negatively correlated with hyperactivity/impulsivity (*β=*-0.35, *p=*0.012) in the comparison sample, but not the at-risk sample (*p*’s>0.355; see Figure S2).

## Discussion

In the current study we tested whether the triple network model, which implicates divergent connectivity between the SN, CEN, and DMN in a range of cognitive and affective conditions, can characterize behavioral difficulties in a broad transdiagnostic sample of children. Hyperactivity/impulsivity across this sample of neurodevelopmentally at-risk children was associated with reduced segregation between the SN, CEN, and DMN. We replicated these effects using networks and regions derived from an adult parcellation of brain function. These findings directly provide transdiagnostic evidence for the triple network model in a neurodevelopmental sample. Interestingly, these associations differed in a sample of comparison children, particularly between the CEN-DMN and within the CEN, suggesting different neurodevelopmental origins. Finally, we replicated our findings when excluding children who moved in the scanner at increasingly stringent thresholds.

### Transdiagnostic evidence for the Triple Network Model in Neurodevelopmental Risk

We found novel evidence for the triple network model (7) in a mixed sample of children with diverse neurodevelopmental characteristics and diagnoses. Importantly, we showed that hyperactivity/impulsivity was associated with higher functional connectivity between all three networks in neurodevelopmentally at-risk children. Larger deviations in functional connectivity were related to a greater degree of neurodevelopmental difficulties, regardless of diagnostic status. In contrast, previous case-control studies have implied that deviations in connectivity are related to the specific symptoms of ADHD or autism (19,20). Our findings were robust to different parcellation methods; we demonstrated that hyperactivity/impulsivity was also associated with greater CEN-DMN and SN-DMN connectivity in the at-risk sample when using regions and networks derived from an adult atlas of functional connectivity. Interestingly, we found less evidence for group differences in connectivity: only SN-CEN functional connectivity was significantly elevated in those diagnosed with combined type ADHD. One possible reason for this may be the substantial heterogeneity in the types and degree of neurodevelopmental difficulties in our at-risk sample (27) and children with ADHD (4), which was better captured by the dimensional analyses. In short, hyperactivity/impulsivity was related to elevated connectivity between the SN, CEN and DMN. But, why might this elevated connectivity occur in neurodevelopment?

### Functional Segregation May underlie Neurodevelopmental Difficulties

The developmental process of functional segregation may diverge in children experiencing neurodevelopmental difficulties. In childhood, brain function increasingly segregates in local neighborhoods and integrates at the global level through networks of coordinated regions (49,50). This phenomenon leads to decreasing functional connectivity between the SN, CEN and DMN through childhood and adolescence (10,11,51–53). The elevated connectivity in hyperactive/impulsive individuals would therefore be similar to younger individuals without these difficulties. Indeed, previous evidence in this at-risk cohort demonstrated that the CEN and a limbic network, which overlaps with the SN component in the mOFC, did not segregate with age relative to comparison children (34). Exploratory analyses in the current study also suggested that the effects of age on CEN-DMN functional connectivity differed between the at-risk and comparison samples. Where the development of these networks is attenuated or delayed, it may act as a particular risk factor for hyperactivity/impulsivity, consistent with evidence in children with a formal ADHD diagnosis (54,55).

Reduced CEN-DMN segregation and reduced CEN integration in children with the most marked hyperactivity/impulsivity may stem from a history of difficulties sustaining CEN activity. The CEN-DMN finding was evidenced in the data-driven networks, adult networks, and between specific regions. It is consistent with many previous studies implicating higher CEN-DMN connectivity in neurodevelopmental difficulties, including hyperactivity/impulsivity and inattention (56–58), IQ (11,52), and inhibitory control (59). The CEN regulates cognitive control by coordinating externally oriented networks and suppressing the internally oriented default-mode network (12). Higher resting CEN-DMN functional connectivity has been implicated in lapses of attention and may reflect a history of difficulties maintaining CEN activity and suppressing DMN activity during attention demanding tasks (Sonuga-Barke, 2007; Castellanos, 2008). Extending this theory, difficulties sustaining CEN activity may also result in failures to inhibit inappropriate behaviors (60), leading to impulsive actions and increased motor activity. Although we found no significant association with inattention, others have (19,57) and we suspect that this is due to a ceiling effect on this specific measure in our sample.

Relatedly, a greater propensity for impulsive behavior in at-risk children could originate from a heightened developmental mismatch between the SN and CEN. The developmental mismatch hypothesis explains how increased sensation seeking and risk-taking behavior in adolescence may arise from the protracted development of cognitive control in the CEN, relative to the early development of affect and reward processing in limbic and subcortical regions, such as the medial OFC and nucleus accumbens (61–64). Notably, these regions were only included in the data-driven SN component, where SN-CEN connectivity was associated with hyperactivity/impulsivity. Greater connectivity between the SN and CEN components is consistent with the idea that regions involved in emotion and reward processing are under-regulated by the CEN, which is in turn linked to impulsive behavior (65). Therefore, children with the greatest neurodevelopmental difficulties could have a heightened developmental mismatch.

### Diverging Brain-Behavior Associations

A consistent and surprising finding was that the at-risk and comparison samples had opposing associations between hyperactivity/impulsivity and CEN-DMN connectivity. As resting-state connectivity likely develops through interactive specialization that is dependent on co-activity between regions (66), then network interactions can adaptively develop through different means and may underlie the distinct associations we observed in the two samples. This was demonstrated in a recent study where CEN-DMN connectivity was positively or negatively related to cognitive ability depending on whether the sample of children were socioeconomically at-risk or not (52). Similarly, while greater CEN-DMN connectivity may reflect difficulties inhibiting inappropriate behaviors in neurodevelopmentally at-risk children, it could be cognitively advantageous in comparison children. In support of this, our exploratory analyses revealed that fluid intelligence was associated with greater CEN-DMN connectivity and less hyperactivity/impulsivity in comparison children, but not at-risk children. This higher CEN-DMN connectivity could be cognitively advantageous for children who rely more on typical DMN functions, such as autobiographical memory and future planning, in cognitively demanding situations (67). Alternatively, typically developing children may rely more upon specialized regions to inhibit DMN activity in situations requiring externally oriented cognitive control (57), inferring that the CEN and DMN can be more regularly co-activated in these children. It is important to note that even the highest hyperactivity/impulsivity score in comparison children was below the average hyperactivity/impulsivity score in at-risk children. Consequently, our finding may reflect that the true relationship between CEN-DMN connectivity and hyperactivity/impulsivity across the samples is non-linear.

### Limitations and Future Directions

We would like to highlight several limitations and future directions for our work. First, our findings pertained to only one measure of hyperactivity/impulsivity and future studies should address the specificity or generality to other neurodevelopmental difficulties. We suspect that this association was not specific and that the measure captured neurodevelopmental difficulties, such as behavioral regulation, more broadly. Second, we note the well-known limitations of self-report data but emphasize that these have clinical utility in the assessment of neurodevelopmental difficulties. Third, we extracted network components by performing a group ICA jointly across both samples. This practically enabled the direct comparison of brain coordinates across individuals, but it does not take into account spatial variation in functional networks within individuals or within samples.

### Conclusion

We demonstrate that the triple network model differentiates behavioral difficulties across a transdiagnostic neurodevelopmental cohort. Specifically, we observed that functional segregation between these networks was generally attenuated in those with the greatest neurodevelopmental difficulties. We suggest multiple mechanisms that may contribute to this, including: delayed development of functional networks, a history of difficulties maintaining CEN activity, and a heightened developmental mismatch between neural systems responsible for cognitive control compared to those for reward/affect processing.

## Data Availability

The ethics approval for the cohort data used does not currently permit open data access. However, the analysis code is available at: https://osf.io/7mptz/ (DOI: 10.17605/OSF.IO/7MPTZ).

## Acknowledgements and Disclosures

The authors were supported by the Medical Research Council program grant MC-A0606-5PQ41. We would like to thank all members of the CALM Team for their help with recruitment, data collection, and data management, as well as all of the children and parents for their participation in the study. The CALM Team includes lead investigators Duncan Astle, Kate Baker, Susan Gathercole, Joni Holmes, Rogier Kievit and Tom Manly. Data collection is assisted by a team of researchers and PhD students that includes Danyal Akarca, Joe Bathelt, Marc Bennett, Madalena Bettencourt, Giacomo Bignardi, Sarah Bishop, Erica Bottacin, Lara Bridge, Diandra Brkic, Annie Bryant, Sally Butterfield, Elizabeth Byrne, Gemma Crickmore, Edwin Dalmaijer, Fánchea Daly, Tina Emery, Laura Forde, Grace Franckel, Delia Furhmann, Andrew Gadie, Sara Gharooni, Jacalyn Guy, Erin Hawkins, Agnieszka Jaroslawska, Sara Joeghan, Amy Johnson, Jonathan Jones, Silvana Mareva, Elise Ng-Cordell, Sinead O’Brien, Cliodhna O’Leary, Joseph Rennie, Ivan Simpson-Kent, Roma Siugzdaite, Tess Smith, Stepheni Uh, Maria Vedechkina, Francesca Woolgar, Natalia Zdorovtsova, and Mengya Zhang. The authors wish to thank the many professionals working in children’s services in the South-East and East of England for their support, and to the children and their families for giving up their time to visit the clinic. We would also like to thank the radiographers who support the excellent pediatric scanning at the MRC CBSU.

The authors report no biomedical financial interests or potential conflicts of interest.

For the purpose of open access, the author has applied a Creative Commons Attribution (CC-BY) licence to any Author Accepted Manuscript version arising from this submission.

## Notes

### Competing Interest Statement

The authors have declared no competing interest.

### Author Declarations

Research Ethics Committee of the National Health Service gave ethical approval for this work. The Centre for Attention Learning and Memory Management Committee of the MRC Cognition and Brain Science Unit gave ethical approval for this work

