## Supplementary materials for "Testing the Triple Network Model of Psychopathology in a Transdiagnostic Neurodevelopmental Cohort"

**Table S1.**

**Triple Network Connectivity and Neurodevelopmental Difficulties in At-Risk Children**

|  | **Hyperactivity/Impulsivity** | | | | | | | **Inattention** | | | | | | |
| --- | --- | --- | --- | --- | --- | --- | --- | --- | --- | --- | --- | --- | --- | --- |
|  | *B* | | *SE* | | 95% CIs | *p* | | *B* | | *SE* | | 95% CIs | | *p* |
| **SN - CEN** | 0.21 | 0.08 | | 0.06:0.36 | | 0.006** | 0.02 | | 0.08 | | -0.14:0.18 | | 0.784 | |
| **SN - DMN** | 0.2 | 0.08 | | 0.04:0.36 | | 0.013* | 0.05 | | 0.09 | | -0.11:0.22 | | 0.525 | |
| **CEN - DMN** | 0.16 | 0.08 | | 0.01:0.32 | | 0.042* | 0.06 | | 0.08 | | -0.11:0.23 | | 0.484 | |

*Note.* Standardised beta coefficients are reported with Standard Errors (SE) and 95% Confidence Intervals (CIs), controlling for age, age^2^, gender, and in-scanner motion. Salience Network (SN), Central Executive Network (CEN), and Default Mode Network (DMN). **p* < 0.05, ***p* < 0.01

**Table S2.**

**Triple Network Connectivity and Neurodevelopmental Difficulties in Comparison Children**

|  | **Hyperactivity/Impulsivity** | | | | **Inattention** | | | | |
| --- | --- | --- | --- | --- | --- | --- | --- | --- | --- |
|  | *B* | *SE* | 95% CIs | *p* | | *B* | *SE* | 95% CIs | *p* |
| **SN - CEN** | 0.04 | 0.14 | -0.25:0.33 | 0.785 | | -0.02 | 0.14 | -0.30:0.26 | 0.9 |
| **SN - DMN** | 0.07 | 0.16 | -0.25:0.38 | 0.669 | | 0.06 | 0.15 | -0.24:0.36 | 0.698 |
| **CEN - DMN** | -0.54 | 0.12 | -0.79:-0.29 | 6.7E-5*** | | -0.12 | 0.14 | -0.40:0.17 | 0.413 |

*Note.* Standardised beta coefficients are reported with Standard Errors (SE) and 95% Confidence Intervals (CIs), controlling for age, age^2^, gender, and in-scanner motion. Salience Network (SN), Central Executive Network (CEN), and Default Mode Network (DMN). ****p* < 0.001

|  | **Hyperactivity/Impulsivity** | | | | **Inattention** | | | | |
| --- | --- | --- | --- | --- | --- | --- | --- | --- | --- |
|  | *B* | *SE* | 95% CIs | *p* | | *B* | *SE* | 95% CIs | *p* |
| **Ventral SN - CEN** | 0.03 | 0.08 | -0.13:0.19 | 0.723 | | 0.02 | 0.09 | -0.15:0.20 | 0.798 |
| **Ventral SN - DMN** | 0.2 | 0.08 | 0.05:0.36 | 0.009** | | 0.03 | 0.08 | -0.13:0.19 | 0.718 |
| **Dorsal SN - CEN** | -0.07 | 0.08 | -0.23:0.09 | 0.373 | | 0.14 | 0.08 | -0.04:0.29 | 0.128 |
| **Dorsal SN - DMN** | 0.15 | 0.08 | 0.00:0.31 | 0.05* | | -0.01 | 0.08 | -0.17:0.15 | 0.902 |
| **CEN - DMN** | 0.21 | 0.08 | 0.05:0.36 | 0.009** | | 0.11 | 0.08 | -0.05:0.27 | 0.189 |

**Table S3.**

**Adult Network Connectivity and Neurodevelopmental Difficulties in At-Risk Children**

*Note.* Standardised beta coefficients are reported with Standard Errors (SE) and 95% Confidence Intervals (CIs), controlling for age, age^2^, gender, and in-scanner motion. Salience Network (SN), Central Executive Network (CEN), and Default Mode Network (DMN). **p* < 0.05, **p* < 0.01

**Table S4.**

**Adult Network Connectivity and Neurodevelopmental Difficulties in Comparison Children**

|  | **Hyperactivity/Impulsivity** | | | | **Inattention** | | | | |
| --- | --- | --- | --- | --- | --- | --- | --- | --- | --- |
|  | *B* | *SE* | 95% CIs | *p* | | *B* | *SE* | 95% CIs | *p* |
| **Ventral SN - CEN** | 0.15 | 0.15 | -0.15:0.46 | 0.317 | | 0 | 0.16 | -0.31:0.31 | 0.998 |
| **Ventral SN - DMN** | -0.16 | 0.14 | -0.45:0.13 | 0.281 | | -0.13 | 0.14 | -0.41:0.16 | 0.385 |
| **Dorsal SN - CEN** | 0.14 | 0.15 | -0.15:0.44 | 0.334 | | 0.07 | 0.14 | -0.22:0.36 | 0.63 |
| **Dorsal SN - DMN** | -0.09 | 0.14 | -0.39:0.20 | 0.515 | | -0.09 | 0.14 | -0.38:0.19 | 0.506 |
| **CEN - DMN** | -0.22 | 0.14 | -0.50:0.05 | 0.11 | | -0.18 | 0.14 | -0.45:0.10 | 0.196 |

*Note.* Standardised beta coefficients are reported with Standard Errors (SE) and 95% Confidence Intervals (CIs), controlling for age, age^2^, gender, and in-scanner motion. Salience Network (SN), Central Executive Network (CEN), and Default Mode Network (DMN).

**Table S5.**

**Network Connectivity Differences between the Samples**

|  | At-Risk | Comparison | t-test | | | Regression | | | |
| --- | --- | --- | --- | --- | --- | --- | --- | --- | --- |
|  | *M* (*SD*) | *M* (*SD*) | *t* | *D* | *p* | *β* | *SE* | CI | *p* |
| **SN - CEN** | 0.11 (0.17) | 0.06 (0.16) | 2.1 | -0.32 | 0.036* | 0.19 | 0.15 | -0.11:0.49 | 0.22 |
| **SN - DMN** | 0.17 (0.19) | 0.19 (0.21) | -0.73 | 0.11 | 0.465 | -0.08 | 0.16 | -0.40:0.23 | 0.599 |
| **CEN - DMN** | 0.19 (0.15) | 0.17 (0.16) | 0.78 | -0.12 | 0.434 | 0.02 | 0.16 | -0.28:0.33 | 0.874 |

*Note.* Standardised beta coefficients are reported with Standard Errors (SE) and 95% Confidence Intervals (CIs), controlling for age, age^2^, gender, and in-scanner motion. Cohen’s D (*D*), Salience Network (SN), Central Executive Network (CEN), Default Mode Network (DMN). **p* < 0.05

**Table S6.**

**Network Connectivity Differences in Children Diagnosed with Combined Type ADHD**

|  | ADHD | Comparison | t-test | | | Regression | | | |
| --- | --- | --- | --- | --- | --- | --- | --- | --- | --- |
|  | *M* (*SD*) | *M* (*SD*) | *t* | *D* | *p* | *β* | *SE* | CI | *p* |
| **SN - CEN** | 0.16 (0.13) | 0.06 (0.16) | 3.08 | 0.7 | 0.003** | 0.56 | 0.24 | 0.08:-1.04 | 0.023* |
| **SN - DMN** | 0.23 (0.17) | 0.19 (0.21) | 0.78 | 0.18 | 0.44 | 0.15 | 0.26 | -0.36:0.67 | 0.554 |
| **CEN - DMN** | 0.24 (0.08) | 0.18 (0.16) | 1.73 | 0.45 | 0.087 | 0.18 | 0.27 | -0.36:0.71 | 0.512 |

*Note.* Standardised beta coefficients are reported with Standard Errors (SE) and 95% Confidence Intervals (CIs), controlling for age, age^2^, gender, and in-scanner motion. Attention Deficit Hyperactivity Disorder (ADHD), Cohen’s D (*D*), Salience Network (SN), Central Executive Network (CEN), Default Mode Network (DMN). **p* < 0.05, ***p* < 0.01

**Table S7.**

**Regional Salience - Central Executive Network Connectivity and Neurodevelopmental Difficulties in At-Risk Children**

|  | **Hyperactivity/Impulsivity** | | | | **Inattention** | | | | |
| --- | --- | --- | --- | --- | --- | --- | --- | --- | --- |
|  | *B* | *SE* | 95% CIs | *p* | | *B* | *SE* | 95% CIs | *p* |
| L SN dAI - L CEN lPFC | 0.06 | 0.08 | -0.10:0.22 | 0.831 | | 0.09 | 0.08 | -0.07:0.26 | 0.787 |
| L SN dAI - L CEN IPS | 0.02 | 0.08 | -0.14:0.18 | 0.919 | | 0.01 | 0.08 | -0.15:0.18 | 0.994 |
| L SN dAI - R CEN lPFC | 0.03 | 0.08 | -0.13:0.19 | 0.868 | | 0 | 0.08 | -0.16:0.16 | 0.994 |
| L SN dAI - R CEN IPS | -0.01 | 0.08 | -0.17:0.15 | 0.919 | | 0.06 | 0.09 | -0.11:0.23 | 0.787 |
| L SN dAC - L CEN lPFC | -0.03 | 0.08 | -0.19:0.13 | 0.868 | | 0.05 | 0.08 | -0.12:0.21 | 0.787 |
| L SN dAC - L CEN IPS | 0.06 | 0.08 | -0.11:0.22 | 0.831 | | 0.11 | 0.08 | -0.06:0.28 | 0.787 |
| L SN dAC - R CEN lPFC | 0.04 | 0.08 | -0.12:0.20 | 0.868 | | 0.11 | 0.08 | -0.06:0.28 | 0.787 |
| L SN dAC - R CEN IPS | -0.06 | 0.08 | -0.22:0.10 | 0.831 | | 0.15 | 0.08 | -0.01:0.31 | 0.787 |
| L SN vAI - L CEN lPFC | 0.11 | 0.08 | -0.04:0.27 | 0.689 | | -0.14 | 0.08 | -0.30:0.03 | 0.787 |
| L SN vAI - L CEN IPS | 0.07 | 0.08 | -0.09:0.23 | 0.831 | | -0.08 | 0.09 | -0.25:0.09 | 0.787 |
| L SN vAI - R CEN lPFC | 0.12 | 0.08 | -0.04:0.28 | 0.689 | | 0 | 0.08 | -0.16:0.16 | 0.994 |
| L SN vAI - R CEN IPS | 0.11 | 0.08 | -0.05:0.27 | 0.689 | | -0.1 | 0.08 | -0.27:0.07 | 0.787 |
| R SN dAI - L CEN lPFC | 0.2 | 0.08 | 0.04:0.36 | 0.394 | | 0.01 | 0.09 | -0.16:0.18 | 0.994 |
| R SN dAI - L CEN IPS | -0.03 | 0.08 | -0.20:0.14 | 0.868 | | 0.08 | 0.09 | -0.09:0.25 | 0.787 |
| R SN dAI - R CEN lPFC | -0.01 | 0.08 | -0.17:0.15 | 0.919 | | -0.05 | 0.08 | -0.22:0.11 | 0.787 |
| R SN dAI - R CEN IPS | -0.07 | 0.08 | -0.24:0.09 | 0.831 | | 0.05 | 0.08 | -0.11:0.22 | 0.787 |
| R SN dAC - L CEN lPFC | -0.04 | 0.08 | -0.20:0.12 | 0.868 | | 0.06 | 0.09 | -0.11:0.23 | 0.787 |
| R SN dAC - L CEN IPS | 0.01 | 0.08 | -0.15:0.17 | 0.919 | | 0.09 | 0.08 | -0.07:0.26 | 0.787 |
| R SN dAC - R CEN lPFC | 0.03 | 0.08 | -0.13:0.19 | 0.868 | | 0.06 | 0.08 | -0.10:0.23 | 0.787 |
| R SN dAC - R CEN IPS | -0.06 | 0.08 | -0.22:0.11 | 0.831 | | 0.05 | 0.09 | -0.12:0.22 | 0.787 |
| R SN vAI - L CEN lPFC | 0.1 | 0.08 | -0.06:0.25 | 0.725 | | 0.01 | 0.08 | -0.16:0.17 | 0.994 |
| R SN vAI - L CEN IPS | 0.11 | 0.08 | -0.05:0.27 | 0.689 | | 0.08 | 0.08 | -0.09:0.24 | 0.787 |
| R SN vAI - R CEN lPFC | 0.11 | 0.08 | -0.05:0.27 | 0.689 | | 0.05 | 0.08 | -0.12:0.21 | 0.787 |
| R SN vAI - R CEN IPS | 0.09 | 0.08 | -0.06:0.25 | 0.725 | | 0.04 | 0.08 | -0.13:0.21 | 0.792 |

*Note.* Standardised beta coefficients are reported with Standard Errors (SE) and 95% Confidence Intervals (CIs), controlling for age, age^2^, gender, and in-scanner motion. Salience Network (SN), Central Executive Network (CEN), Left (L), Right (R), dorsal Anterior Insula (dAI), dorsal Anterior Cingulate (dAC), ventral Anterior Insula (vAI), lateral Prefrontal Cortex (lPFC), and Intraparietal Sulcus (IPS).

**Table S8.**

**Regional Salience - Central Executive Network Connectivity and Neurodevelopmental Difficulties in Comparison Children**

*Note.* Standardised beta coefficients are reported with Standard Errors (SE) and 95% Confidence Intervals (CIs), controlling for age, age^2^, gender, and in-scanner motion. Left (L), Right (R), Salience Network (SN), Central Executive Network (CEN), dorsal Anterior Insula (dAI), dorsal Anterior Cingulate (dAC), ventral Anterior Insula (vAI), lateral Prefrontal Cortex (lPFC), and Intraparietal Sulcus (IPS).

|  | **Hyperactivity/Impulsivity** | | | | **Inattention** | | | | |
| --- | --- | --- | --- | --- | --- | --- | --- | --- | --- |
|  | *B* | *SE* | 95% CIs | *p* | | *B* | *SE* | 95% CIs | *p* |
| L SN dAI - L CEN lPFC | 0.26 | 0.14 | -0.02:0.55 | 0.324 | | 0.1 | 0.14 | -0.19:0.39 | 0.755 |
| L SN dAI - L CEN IPS | 0.14 | 0.15 | -0.16:0.43 | 0.751 | | 0.11 | 0.14 | -0.18:0.39 | 0.755 |
| L SN dAI - R CEN lPFC | 0.08 | 0.14 | -0.21:0.36 | 0.834 | | 0.09 | 0.14 | -0.18:0.37 | 0.755 |
| L SN dAI - R CEN IPS | 0.06 | 0.15 | -0.25:0.37 | 0.877 | | 0.17 | 0.15 | -0.13:0.48 | 0.755 |
| L SN dAC - L CEN lPFC | -0.18 | 0.15 | -0.48:0.12 | 0.685 | | -0.03 | 0.15 | -0.34:0.28 | 0.963 |
| L SN dAC - L CEN IPS | 0.02 | 0.15 | -0.28:0.32 | 0.961 | | 0.09 | 0.15 | -0.21:0.39 | 0.755 |
| L SN dAC - R CEN lPFC | -0.02 | 0.15 | -0.31:0.27 | 0.961 | | -0.01 | 0.15 | -0.30:0.28 | 0.996 |
| L SN dAC - R CEN IPS | -0.01 | 0.15 | -0.31:0.30 | 0.961 | | 0.18 | 0.14 | -0.11:0.47 | 0.755 |
| L SN vAI - L CEN lPFC | 0.03 | 0.14 | -0.26:0.32 | 0.961 | | 0.08 | 0.14 | -0.20:0.37 | 0.755 |
| L SN vAI - L CEN IPS | -0.11 | 0.14 | -0.40:0.18 | 0.821 | | -0.13 | 0.14 | -0.41:0.15 | 0.755 |
| L SN vAI - R CEN lPFC | -0.26 | 0.15 | -0.55:0.03 | 0.324 | | -0.24 | 0.15 | -0.53:0.05 | 0.755 |
| L SN vAI - R CEN IPS | -0.29 | 0.15 | -0.59:0.01 | 0.324 | | -0.09 | 0.15 | -0.39:0.20 | 0.755 |
| R SN dAI - L CEN lPFC | 0.34 | 0.14 | 0.05:0.62 | 0.324 | | 0.1 | 0.14 | -0.19:0.39 | 0.755 |
| R SN dAI - L CEN IPS | 0.1 | 0.15 | -0.21:0.40 | 0.821 | | 0 | 0.15 | -0.29:0.30 | 0.996 |
| R SN dAI - R CEN lPFC | 0.13 | 0.15 | -0.16:0.43 | 0.751 | | 0.09 | 0.14 | -0.20:0.38 | 0.755 |
| R SN dAI - R CEN IPS | 0.16 | 0.15 | -0.14:0.45 | 0.751 | | 0.07 | 0.15 | -0.23:0.37 | 0.806 |
| R SN dAC - L CEN lPFC | -0.01 | 0.15 | -0.32:0.29 | 0.961 | | -0.15 | 0.15 | -0.45:0.15 | 0.755 |
| R SN dAC - L CEN IPS | 0.1 | 0.16 | -0.23:0.43 | 0.821 | | -0.15 | 0.16 | -0.47:0.16 | 0.755 |
| R SN dAC - R CEN lPFC | -0.06 | 0.15 | -0.37:0.25 | 0.877 | | -0.13 | 0.15 | -0.43:0.16 | 0.755 |
| R SN dAC - R CEN IPS | 0.19 | 0.15 | -0.11:0.50 | 0.685 | | 0.02 | 0.15 | -0.27:0.31 | 0.963 |
| R SN vAI - L CEN lPFC | 0.26 | 0.14 | -0.03:0.54 | 0.324 | | 0.15 | 0.14 | -0.14:0.43 | 0.755 |
| R SN vAI - L CEN IPS | 0.31 | 0.13 | 0.04:0.58 | 0.324 | | -0.04 | 0.14 | -0.31:0.24 | 0.959 |
| R SN vAI - R CEN lPFC | -0.15 | 0.17 | -0.49:0.19 | 0.751 | | -0.19 | 0.15 | -0.49:0.12 | 0.755 |
| R SN vAI - R CEN IPS | 0.09 | 0.15 | -0.21:0.40 | 0.821 | | -0.09 | 0.15 | -0.39:0.20 | 0.755 |

**Table S9.**

**Regional Salience – Default Mode Network Connectivity and Neurodevelopmental Difficulties in At-Risk Children**

|  | **Hyperactivity/Impulsivity** | | | | **Inattention** | | | | |
| --- | --- | --- | --- | --- | --- | --- | --- | --- | --- |
|  | *B* | *SE* | 95% CIs | *p* | | *B* | *SE* | 95% CIs | *p* |
| L SN dAI - L DMN mPFC | -0.02 | 0.08 | -0.17:0.13 | 0.838 | | 0.12 | 0.08 | -0.04:0.27 | 0.834 |
| L SN dAI - L DMN PCC | 0.07 | 0.08 | -0.08:0.23 | 0.482 | | 0.02 | 0.08 | -0.14:0.18 | 0.856 |
| L SN dAI - R DMN mPFC | 0.05 | 0.08 | -0.10:0.20 | 0.593 | | 0.14 | 0.08 | -0.01:0.30 | 0.834 |
| L SN dAI - R DMN PCC | 0.1 | 0.08 | -0.06:0.26 | 0.412 | | 0.04 | 0.08 | -0.13:0.20 | 0.834 |
| L SN dAC - L DMN mPFC | 0.08 | 0.08 | -0.08:0.24 | 0.482 | | 0.03 | 0.09 | -0.14:0.20 | 0.852 |
| L SN dAC - L DMN PCC | 0.18 | 0.08 | 0.02:0.34 | 0.125 | | -0.07 | 0.08 | -0.24:0.09 | 0.834 |
| L SN dAC - R DMN mPFC | 0.12 | 0.08 | -0.04:0.28 | 0.351 | | -0.08 | 0.08 | -0.25:0.08 | 0.834 |
| L SN dAC - R DMN PCC | 0.16 | 0.08 | 0.00:0.32 | 0.149 | | -0.09 | 0.08 | -0.26:0.07 | 0.834 |
| L SN vAI - L DMN mPFC | 0 | 0.08 | -0.16:0.17 | 0.985 | | 0.02 | 0.08 | -0.15:0.19 | 0.856 |
| L SN vAI - L DMN PCC | 0.17 | 0.08 | 0.01:0.32 | 0.139 | | 0.05 | 0.08 | -0.11:0.21 | 0.834 |
| L SN vAI - R DMN mPFC | 0.06 | 0.08 | -0.10:0.22 | 0.565 | | 0.04 | 0.08 | -0.13:0.21 | 0.834 |
| L SN vAI - R DMN PCC | 0.13 | 0.08 | -0.03:0.29 | 0.337 | | 0.06 | 0.08 | -0.11:0.23 | 0.834 |
| R SN dAI - L DMN mPFC | 0 | 0.08 | -0.16:0.15 | 0.985 | | 0.06 | 0.08 | -0.10:0.22 | 0.834 |
| R SN dAI - L DMN PCC | 0.12 | 0.08 | -0.04:0.28 | 0.337 | | -0.05 | 0.08 | -0.22:0.11 | 0.834 |
| R SN dAI - R DMN mPFC | 0.07 | 0.08 | -0.09:0.23 | 0.499 | | 0.08 | 0.08 | -0.08:0.25 | 0.834 |
| R SN dAI - R DMN PCC | 0.07 | 0.08 | -0.08:0.23 | 0.482 | | -0.07 | 0.08 | -0.23:0.09 | 0.834 |
| R SN dAC - L DMN mPFC | 0.09 | 0.08 | -0.07:0.24 | 0.435 | | 0.09 | 0.08 | -0.07:0.25 | 0.834 |
| R SN dAC - L DMN PCC | 0.26 | 0.08 | 0.10:0.41 | 0.028* | | 0.05 | 0.08 | -0.12:0.21 | 0.834 |
| R SN dAC - R DMN mPFC | 0.09 | 0.08 | -0.06:0.25 | 0.412 | | 0.05 | 0.08 | -0.12:0.21 | 0.834 |
| R SN dAC - R DMN PCC | 0.22 | 0.08 | 0.06:0.37 | 0.061 | | -0.04 | 0.08 | -0.20:0.13 | 0.834 |
| R SN vAI - L DMN mPFC | 0.1 | 0.08 | -0.06:0.26 | 0.412 | | 0.03 | 0.08 | -0.14:0.19 | 0.856 |
| R SN vAI - L DMN PCC | 0.21 | 0.08 | 0.05:0.37 | 0.061^+^ | | 0.05 | 0.09 | -0.12:0.22 | 0.834 |
| R SN vAI - R DMN mPFC | 0.1 | 0.08 | -0.05:0.26 | 0.412 | | -0.02 | 0.08 | -0.18:0.15 | 0.856 |
| R SN vAI - R DMN PCC | 0.21 | 0.08 | 0.06:0.36 | 0.061^+^ | | 0.04 | 0.08 | -0.12:0.20 | 0.834 |

*Note.* Standardised beta coefficients are reported with Standard Errors (SE) and 95% Confidence Intervals (CIs), controlling for age, age^2^, gender, and in-scanner motion. Left (L), Right (R), Salience Network (SN), Default Mode Network (DMN), dorsal Anterior Insula (dAI), dorsal Anterior Cingulate (dAC), ventral Anterior Insula (vAI), medial Prefrontal Cortex (mPFC), and Posterior Cingulate Cortex (PCC). ^+^p < 0.062, *p < 0.05

**Table S10.**

**Regional Salience – Default Mode Network Connectivity and Neurodevelopmental Difficulties in Comparison**

|  | **Hyperactivity/Impulsivity** | | | | **Inattention** | | | | |
| --- | --- | --- | --- | --- | --- | --- | --- | --- | --- |
|  | *B* | *SE* | 95% CIs | *p* | | *B* | *SE* | 95% CIs | *p* |
| L SN dAI - L DMN mPFC | -0.12 | 0.15 | -0.41:0.17 | 0.879 | | -0.05 | 0.14 | -0.34:0.24 | 0.949 |
| L SN dAI - L DMN PCC | 0.11 | 0.15 | -0.18:0.41 | 0.879 | | 0.11 | 0.14 | -0.18:0.40 | 0.949 |
| L SN dAI - R DMN mPFC | -0.18 | 0.15 | -0.48:0.13 | 0.879 | | -0.3 | 0.15 | -0.59:-0.00 | 0.949 |
| L SN dAI - R DMN PCC | 0.06 | 0.14 | -0.23:0.36 | 0.879 | | 0.13 | 0.14 | -0.15:0.41 | 0.949 |
| L SN dAC - L DMN mPFC | -0.08 | 0.15 | -0.37:0.22 | 0.879 | | -0.2 | 0.14 | -0.49:0.08 | 0.949 |
| L SN dAC - L DMN PCC | -0.09 | 0.14 | -0.38:0.20 | 0.879 | | -0.05 | 0.14 | -0.33:0.24 | 0.949 |
| L SN dAC - R DMN mPFC | -0.11 | 0.16 | -0.42:0.20 | 0.879 | | -0.11 | 0.15 | -0.41:0.20 | 0.949 |
| L SN dAC - R DMN PCC | -0.02 | 0.14 | -0.30:0.27 | 0.946 | | -0.03 | 0.14 | -0.31:0.25 | 0.97 |
| L SN vAI - L DMN mPFC | 0.07 | 0.15 | -0.23:0.37 | 0.879 | | 0.01 | 0.15 | -0.29:0.30 | 0.973 |
| L SN vAI - L DMN PCC | 0.09 | 0.15 | -0.22:0.39 | 0.879 | | -0.06 | 0.15 | -0.37:0.24 | 0.949 |
| L SN vAI - R DMN mPFC | -0.05 | 0.14 | -0.33:0.24 | 0.943 | | 0.06 | 0.15 | -0.23:0.35 | 0.949 |
| L SN vAI - R DMN PCC | -0.01 | 0.15 | -0.31:0.29 | 0.946 | | -0.07 | 0.14 | -0.36:0.22 | 0.949 |
| R SN dAI - L DMN mPFC | -0.01 | 0.15 | -0.32:0.30 | 0.946 | | -0.05 | 0.15 | -0.35:0.26 | 0.949 |
| R SN dAI - L DMN PCC | 0.09 | 0.15 | -0.20:0.39 | 0.879 | | 0.07 | 0.15 | -0.22:0.36 | 0.949 |
| R SN dAI - R DMN mPFC | 0.03 | 0.15 | -0.28:0.34 | 0.946 | | -0.1 | 0.15 | -0.40:0.20 | 0.949 |
| R SN dAI - R DMN PCC | 0.12 | 0.15 | -0.19:0.42 | 0.879 | | 0.06 | 0.15 | -0.24:0.35 | 0.949 |
| R SN dAC - L DMN mPFC | -0.18 | 0.14 | -0.47:0.11 | 0.879 | | -0.11 | 0.14 | -0.40:0.17 | 0.949 |
| R SN dAC - L DMN PCC | -0.09 | 0.14 | -0.38:0.19 | 0.879 | | 0.06 | 0.14 | -0.22:0.33 | 0.949 |
| R SN dAC - R DMN mPFC | -0.08 | 0.15 | -0.37:0.22 | 0.879 | | -0.09 | 0.14 | -0.38:0.19 | 0.949 |
| R SN dAC - R DMN PCC | -0.03 | 0.14 | -0.31:0.24 | 0.946 | | -0.01 | 0.13 | -0.28:0.25 | 0.973 |
| R SN vAI - L DMN mPFC | -0.27 | 0.14 | -0.55:0.02 | 0.795 | | 0.02 | 0.14 | -0.27:0.31 | 0.973 |
| R SN vAI - L DMN PCC | -0.23 | 0.15 | -0.54:0.07 | 0.879 | | -0.13 | 0.15 | -0.44:0.18 | 0.949 |
| R SN vAI - R DMN mPFC | -0.35 | 0.14 | -0.64:-0.06 | 0.421 | | -0.01 | 0.15 | -0.31:0.29 | 0.973 |
| R SN vAI - R DMN PCC | -0.14 | 0.15 | -0.45:0.17 | 0.879 | | -0.22 | 0.15 | -0.52:0.08 | 0.949 |

*Note.* Standardised beta coefficients are reported with Standard Errors (SE) and 95% Confidence Intervals (CIs), controlling for age, age^2^, gender, and in-scanner motion. Left (L), Right (R), Salience Network (SN), Default Mode Network (DMN), dorsal Anterior Insula (dAI), dorsal Anterior Cingulate (dAC), ventral Anterior Insula (vAI), medial Prefrontal Cortex (mPFC), and Posterior Cingulate Cortex (PCC).

**Table S11.**

**Regional Central Executive – Default Mode Network Connectivity and Neurodevelopmental Difficulties in At-Risk Children**

|  | **Hyperactivity/Impulsivity** | | | | **Inattention** | | | | |
| --- | --- | --- | --- | --- | --- | --- | --- | --- | --- |
|  | *B* | *SE* | 95% CIs | *p* | | *B* | *SE* | 95% CIs | *p* |
| L DMN mPFC - L CEN lPFC | 0.19 | 0.08 | 0.03:0.35 | 0.072^+^ | | 0.05 | 0.08 | -0.12:0.21 | 0.668 |
| L DMN mPFC - L CEN IPS | 0.18 | 0.08 | 0.03:0.33 | 0.072^+^ | | 0.11 | 0.08 | -0.05:0.27 | 0.58 |
| L DMN mPFC - R CEN lPFC | 0.29 | 0.08 | 0.14:0.43 | 0.003** | | 0.14 | 0.08 | -0.02:0.29 | 0.478 |
| L DMN mPFC - R CEN IPS | 0.06 | 0.08 | -0.10:0.22 | 0.613 | | 0.1 | 0.08 | -0.07:0.26 | 0.58 |
| L DMN PCC - L CEN lPFC | 0.07 | 0.08 | -0.09:0.23 | 0.597 | | 0.07 | 0.09 | -0.10:0.24 | 0.616 |
| L DMN PCC - L CEN IPS | -0.01 | 0.08 | -0.17:0.15 | 0.958 | | 0.04 | 0.08 | -0.13:0.21 | 0.68 |
| L DMN PCC - R CEN lPFC | 0.16 | 0.08 | 0.01:0.32 | 0.108 | | 0.06 | 0.08 | -0.11:0.22 | 0.616 |
| L DMN PCC - R CEN IPS | 0.11 | 0.08 | -0.05:0.27 | 0.367 | | -0.06 | 0.09 | -0.23:0.11 | 0.616 |
| R DMN mPFC - L CEN lPFC | 0.18 | 0.08 | 0.03:0.34 | 0.072^+^ | | 0.02 | 0.08 | -0.14:0.19 | 0.778 |
| R DMN mPFC - L CEN IPS | 0.14 | 0.08 | -0.02:0.29 | 0.184 | | 0.14 | 0.08 | -0.02:0.30 | 0.478 |
| R DMN mPFC - R CEN lPFC | 0.25 | 0.08 | 0.10:0.40 | 0.008** | | 0.07 | 0.08 | -0.09:0.24 | 0.58 |
| R DMN mPFC - R CEN IPS | 0.03 | 0.08 | -0.13:0.19 | 0.886 | | 0.1 | 0.08 | -0.07:0.26 | 0.58 |
| R DMN PCC - L CEN lPFC | 0.07 | 0.08 | -0.10:0.23 | 0.606 | | 0.1 | 0.08 | -0.07:0.27 | 0.58 |
| R DMN PCC - L CEN IPS | 0.02 | 0.08 | -0.14:0.18 | 0.929 | | 0.08 | 0.08 | -0.09:0.24 | 0.58 |
| R DMN PCC - R CEN lPFC | 0.08 | 0.08 | -0.08:0.24 | 0.543 | | 0.09 | 0.08 | -0.08:0.25 | 0.58 |
| R DMN PCC - R CEN IPS | 0 | 0.08 | -0.16:0.17 | 0.959 | | -0.16 | 0.08 | -0.33:0.00 | 0.478 |

*Note.* Standardised beta coefficients are reported with Standard Errors (SE) and 95% Confidence Intervals (CIs), controlling for age, age^2^, gender, and in-scanner motion. Left (L), Right (R), Central Executive Network (CEN), Default Mode Network (DMN), lateral Prefrontal Cortex (lPFC), Intraparietal Sulcus (IPS), medial Prefrontal Cortex (mPFC), and Posterior Cingulate Cortex (PCC). ^+^*p* < 0.073, **p* < 0.05, **p* < 0.01

**Table S12.**

**Regional Central Executive – Default Mode Network Connectivity and Neurodevelopmental Difficulties in Comparison Children**

|  | **Hyperactivity/Impulsivity** | | | | **Inattention** | | | | |
| --- | --- | --- | --- | --- | --- | --- | --- | --- | --- |
|  | *B* | *SE* | 95% CIs | *p* | | *B* | *SE* | 95% CIs | *p* |
| L DMN mPFC - L CEN lPFC | -0.23 | 0.13 | -0.50:0.04 | 0.344 | | -0.02 | 0.14 | -0.30:0.26 | 0.879 |
| L DMN mPFC - L CEN IPS | -0.19 | 0.14 | -0.48:0.09 | 0.344 | | -0.19 | 0.14 | -0.47:0.09 | 0.594 |
| L DMN mPFC - R CEN lPFC | -0.24 | 0.15 | -0.53:0.06 | 0.344 | | -0.16 | 0.15 | -0.46:0.13 | 0.594 |
| L DMN mPFC - R CEN IPS | -0.2 | 0.15 | -0.50:0.09 | 0.344 | | -0.04 | 0.14 | -0.33:0.25 | 0.835 |
| L DMN PCC - L CEN lPFC | -0.28 | 0.15 | -0.58:0.01 | 0.344 | | -0.11 | 0.16 | -0.43:0.20 | 0.666 |
| L DMN PCC - L CEN IPS | -0.08 | 0.14 | -0.37:0.20 | 0.637 | | -0.15 | 0.14 | -0.43:0.13 | 0.594 |
| L DMN PCC - R CEN lPFC | -0.24 | 0.14 | -0.52:0.04 | 0.344 | | -0.17 | 0.14 | -0.44:0.11 | 0.594 |
| L DMN PCC - R CEN IPS | -0.12 | 0.15 | -0.42:0.17 | 0.566 | | 0.07 | 0.14 | -0.22:0.36 | 0.767 |
| R DMN mPFC - L CEN lPFC | -0.15 | 0.13 | -0.41:0.11 | 0.401 | | -0.09 | 0.13 | -0.35:0.17 | 0.666 |
| R DMN mPFC - L CEN IPS | -0.02 | 0.15 | -0.33:0.28 | 0.88 | | -0.16 | 0.15 | -0.45:0.14 | 0.594 |
| R DMN mPFC - R CEN lPFC | -0.22 | 0.15 | -0.53:0.09 | 0.344 | | -0.28 | 0.15 | -0.58:0.02 | 0.594 |
| R DMN mPFC - R CEN IPS | -0.12 | 0.15 | -0.43:0.18 | 0.566 | | -0.21 | 0.15 | -0.50:0.08 | 0.594 |
| R DMN PCC - L CEN lPFC | -0.25 | 0.14 | -0.53:0.03 | 0.344 | | -0.12 | 0.14 | -0.41:0.16 | 0.666 |
| R DMN PCC - L CEN IPS | -0.04 | 0.14 | -0.32:0.24 | 0.852 | | -0.1 | 0.14 | -0.37:0.17 | 0.666 |
| R DMN PCC - R CEN lPFC | -0.18 | 0.14 | -0.46:0.10 | 0.344 | | -0.17 | 0.14 | -0.44:0.10 | 0.594 |
| R DMN PCC - R CEN IPS | -0.09 | 0.15 | -0.38:0.20 | 0.637 | | -0.04 | 0.14 | -0.33:0.25 | 0.835 |

*Note.* Standardised beta coefficients are reported with Standard Errors (SE) and 95% Confidence Intervals (CIs), controlling for age, age^2^, gender, and in-scanner motion. Left (L), Right (R), Central Executive Network (CEN), Default Mode Network (DMN), lateral Prefrontal Cortex (lPFC), Intraparietal Sulcus (IPS), medial Prefrontal Cortex (mPFC), and Posterior Cingulate Cortex (PCC).

**Table S13.**

**Within Salience Network Connectivity and Neurodevelopmental Difficulties in At-Risk Children**

|  | **Hyperactivity/Impulsivity** | | | | **Inattention** | | | | |
| --- | --- | --- | --- | --- | --- | --- | --- | --- | --- |
|  | *B* | *SE* | 95% CIs | *p* | | *B* | *SE* | 95% CIs | *p* |
| L SN dAI - L SN dAC | -0.14 | 0.08 | -0.29:0.02 | 0.418 | | 0.08 | 0.08 | -0.09:0.25 | 0.571 |
| L SN dAI - L SN vAI | -0.1 | 0.08 | -0.26:0.06 | 0.46 | | -0.02 | 0.08 | -0.18:0.14 | 0.878 |
| L SN dAI - R SN AI | 0.01 | 0.08 | -0.15:0.17 | 0.894 | | 0.18 | 0.08 | 0.01:0.34 | 0.351 |
| L SN dAI - R SN dAC | -0.1 | 0.08 | -0.26:0.06 | 0.46 | | -0.01 | 0.09 | -0.18:0.16 | 0.878 |
| L SN dAC - L SN vAI | -0.01 | 0.08 | -0.17:0.15 | 0.894 | | -0.13 | 0.08 | -0.29:0.04 | 0.5 |
| L SN dAC - R SN AI | -0.09 | 0.08 | -0.24:0.07 | 0.46 | | 0.12 | 0.08 | -0.05:0.28 | 0.5 |
| L SN dAC - R SN dAC | -0.01 | 0.08 | -0.17:0.15 | 0.894 | | 0.03 | 0.08 | -0.14:0.20 | 0.878 |
| L SN vAI - R SN AI | -0.1 | 0.08 | -0.26:0.07 | 0.46 | | 0.09 | 0.09 | -0.08:0.25 | 0.571 |
| L SN vAI - R SN dAC | -0.08 | 0.08 | -0.24:0.08 | 0.46 | | -0.11 | 0.08 | -0.28:0.06 | 0.5 |
| R SN dAI - R SN dAC | -0.21 | 0.08 | -0.37:-0.05 | 0.088 | | 0.04 | 0.09 | -0.13:0.21 | 0.878 |

*Note.* Standardised beta coefficients are reported with Standard Errors (SE) and 95% Confidence Intervals (CIs), controlling for age, age^2^, gender, and in-scanner motion. Left (L), Right (R), Salience Network (SN), dorsal Anterior Insula (dAI), dorsal Anterior Cingulate (dAC), and ventral Anterior Insula (vAI).

**Table S14.**

|  | **Hyperactivity/Impulsivity** | | | | **Inattention** | | | | |
| --- | --- | --- | --- | --- | --- | --- | --- | --- | --- |
|  | *B* | *SE* | 95% CIs | *p* | | *B* | *SE* | 95% CIs | *p* |
| L SN dAI - L SN dAC | -0.08 | 0.15 | -0.39:0.22 | 0.922 | | -0.2 | 0.15 | -0.50:0.10 | 0.928 |
| L SN dAI - L SN vAI | -0.1 | 0.15 | -0.39:0.20 | 0.922 | | 0.07 | 0.15 | -0.22:0.37 | 0.928 |
| L SN dAI - R SN AI | -0.04 | 0.15 | -0.34:0.26 | 0.922 | | 0.06 | 0.14 | -0.23:0.35 | 0.928 |
| L SN dAI - R SN dAC | -0.07 | 0.15 | -0.38:0.23 | 0.922 | | 0.01 | 0.15 | -0.29:0.32 | 0.928 |
| L SN dAC - L SN vAI | 0.03 | 0.15 | -0.27:0.33 | 0.922 | | 0.04 | 0.15 | -0.26:0.33 | 0.928 |
| L SN dAC - R SN AI | 0.06 | 0.15 | -0.24:0.36 | 0.922 | | -0.02 | 0.15 | -0.32:0.27 | 0.928 |
| L SN dAC - R SN dAC | -0.19 | 0.15 | -0.50:0.12 | 0.922 | | 0.02 | 0.15 | -0.28:0.32 | 0.928 |
| L SN vAI - R SN AI | 0.01 | 0.15 | -0.28:0.31 | 0.932 | | -0.09 | 0.15 | -0.39:0.21 | 0.928 |
| L SN vAI - R SN dAC | -0.23 | 0.14 | -0.52:0.06 | 0.922 | | -0.28 | 0.14 | -0.56:0.00 | 0.537 |
| R SN dAI - R SN dAC | 0.04 | 0.15 | -0.27:0.34 | 0.922 | | -0.1 | 0.15 | -0.40:0.20 | 0.928 |

**Within Salience Network Connectivity and Neurodevelopmental Difficulties in Comparison Children**

*Note.* Standardised beta coefficients are reported with Standard Errors (SE) and 95% Confidence Intervals (CIs), controlling for age, age^2^, gender, and in-scanner motion. Left (L), Right (R), Salience Network (SN), dorsal Anterior Insula (dAI), dorsal Anterior Cingulate (dAC), and ventral Anterior Insula (vAI).

**Table S15.**

|  | **Hyperactivity/Impulsivity** | | | | **Inattention** | | | | |
| --- | --- | --- | --- | --- | --- | --- | --- | --- | --- |
|  | *B* | *SE* | 95% CIs | *p* | | *B* | *SE* | 95% CIs | *p* |
| L CEN lPFC - L CEN IPS | -0.07 | 0.08 | -0.23:0.09 | 0.468 | | 0.08 | 0.08 | -0.08:0.25 | 0.641 |
| L CEN lPFC - R CEN lPFC | 0.14 | 0.08 | -0.02:0.30 | 0.281 | | 0.07 | 0.09 | -0.10:0.24 | 0.641 |
| L CEN lPFC - R CEN IPS | 0.01 | 0.08 | -0.15:0.17 | 0.926 | | -0.01 | 0.08 | -0.18:0.16 | 0.998 |
| L CEN IPS - R CEN lPFC | -0.09 | 0.08 | -0.25:0.07 | 0.468 | | 0 | 0.09 | -0.17:0.17 | 0.998 |
| L CEN IPS - R CEN IPS | 0.07 | 0.08 | -0.09:0.23 | 0.468 | | -0.15 | 0.08 | -0.32:0.01 | 0.397 |
| R CEN lPFC - R CEN IPS | -0.22 | 0.08 | -0.38:-0.06 | 0.04* | | -0.09 | 0.09 | -0.26:0.08 | 0.641 |

**Within Central Executive Network Connectivity and Neurodevelopmental Difficulties in At-Risk Children**

*Note.* Standardised beta coefficients are reported with Standard Errors (SE) and 95% Confidence Intervals (CIs), controlling for age, age^2^, gender, and in-scanner motion. Left (L), Right (R), Central Executive Network (CEN), lateral Prefrontal Cortex (lPFC), and Intraparietal Sulcus (IPS). **p* < 0.05

**Table S16.**

|  | **Hyperactivity/Impulsivity** | | | | **Inattention** | | | | |
| --- | --- | --- | --- | --- | --- | --- | --- | --- | --- |
|  | *B* | *SE* | 95% CIs | *p* | | *B* | *SE* | 95% CIs | *p* |
| L CEN lPFC - L CEN IPS | -0.35 | 0.13 | -0.62:-0.09 | 0.046* | | -0.02 | 0.13 | -0.29:0.25 | 0.901 |
| L CEN lPFC - R CEN lPFC | 0.15 | 0.16 | -0.18:0.47 | 0.747 | | 0.05 | 0.16 | -0.27:0.37 | 0.901 |
| L CEN lPFC - R CEN IPS | -0.02 | 0.15 | -0.33:0.29 | 0.964 | | -0.02 | 0.15 | -0.31:0.28 | 0.901 |
| L CEN IPS - R CEN lPFC | 0.01 | 0.15 | -0.30:0.31 | 0.964 | | 0.04 | 0.15 | -0.27:0.34 | 0.901 |
| L CEN IPS - R CEN IPS | 0.06 | 0.15 | -0.24:0.35 | 0.964 | | -0.07 | 0.15 | -0.36:0.23 | 0.901 |
| R CEN lPFC - R CEN IPS | 0.36 | 0.14 | 0.07:0.64 | 0.046* | | 0.09 | 0.15 | -0.21:0.39 | 0.901 |

**Within Central Executive Network Connectivity and Neurodevelopmental Difficulties in Comparison Children**

*Note.* Standardised beta coefficients are reported with Standard Errors (SE) and 95% Confidence Intervals (CIs), controlling for age, age^2^, gender, and in-scanner motion. Left (L), Right (R), Central Executive Network (CEN), lateral Prefrontal Cortex (lPFC), and Intraparietal Sulcus (IPS). **p* < 0.05

**Table S17.**

|  | **Hyperactivity/Impulsivity** | | | | **Inattention** | | | | |
| --- | --- | --- | --- | --- | --- | --- | --- | --- | --- |
|  | *B* | *SE* | 95% CIs | *p* | | *B* | *SE* | 95% CIs | *p* |
| L DMN mPFC - L DMN PCC | 0.14 | 0.08 | -0.02:0.30 | 0.162 | | -0.1 | 0.08 | -0.26:0.07 | 0.514 |
| L DMN mPFC - R DMN mPFC | 0.03 | 0.08 | -0.13:0.19 | 0.82 | | 0.03 | 0.08 | -0.14:0.19 | 0.742 |
| L DMN mPFC - R DMN PCC | 0.17 | 0.08 | 0.01:0.33 | 0.102 | | -0.07 | 0.08 | -0.23:0.10 | 0.514 |
| L DMN PCC - R DMN mPFC | 0.22 | 0.08 | 0.06:0.38 | 0.042* | | -0.07 | 0.08 | -0.24:0.10 | 0.514 |
| L DMN PCC - R DMN PCC | 0.02 | 0.08 | -0.14:0.18 | 0.82 | | -0.11 | 0.08 | -0.28:0.05 | 0.514 |
| R DMN mPFC - R DMN PCC | 0.11 | 0.08 | -0.04:0.27 | 0.237 | | -0.08 | 0.08 | -0.24:0.08 | 0.514 |

**Within Default Mode Network Connectivity and Neurodevelopmental Difficulties in At-Risk Children**

*Note.* Standardised beta coefficients are reported with Standard Errors (SE) and 95% Confidence Intervals (CIs), controlling for age, age^2^, gender, and in-scanner motion. Left (L), Right (R), Default Mode Network (DMN), medial Prefrontal Cortex (mPFC), and Posterior Cingulate Cortex (PCC). **p* < 0.05

**Table S18.**

|  | **Hyperactivity/Impulsivity** | | | | **Inattention** | | | | |
| --- | --- | --- | --- | --- | --- | --- | --- | --- | --- |
|  | *B* | *SE* | 95% CIs | *p* | | *B* | *SE* | 95% CIs | *p* |
| L DMN mPFC - L DMN PCC | 0.03 | 0.15 | -0.28:0.34 | 0.845 | | -0.04 | 0.15 | -0.34:0.26 | 0.993 |
| L DMN mPFC - R DMN mPFC | 0.07 | 0.15 | -0.22:0.36 | 0.845 | | -0.01 | 0.14 | -0.30:0.27 | 0.993 |
| L DMN mPFC - R DMN PCC | 0.08 | 0.16 | -0.23:0.39 | 0.845 | | 0 | 0.15 | -0.31:0.30 | 0.993 |
| L DMN PCC - R DMN mPFC | -0.17 | 0.15 | -0.48:0.13 | 0.845 | | -0.09 | 0.15 | -0.39:0.21 | 0.993 |
| L DMN PCC - R DMN PCC | -0.1 | 0.16 | -0.41:0.22 | 0.845 | | 0.02 | 0.15 | -0.28:0.32 | 0.993 |
| R DMN mPFC - R DMN PCC | -0.06 | 0.16 | -0.37:0.26 | 0.845 | | -0.04 | 0.16 | -0.35:0.28 | 0.993 |

**Within Default Mode Network Connectivity and Neurodevelopmental Difficulties in Comparison Children**

*Note.* Standardised beta coefficients are reported with Standard Errors (SE) and 95% Confidence Intervals (CIs), controlling for age, age^2^, gender, and in-scanner motion. Left (L), Right (R), Default Mode Network (DMN), medial Prefrontal Cortex (mPFC), and Posterior Cingulate Cortex (PCC).

**Table S19**

**Triple Network Connectivity and Neurodevelopmental Difficulties in At-Risk Children across Motion Thresholds**

|  | **Hyperactivity/Impulsivity** | | | | | | | **Inattention** | | | | |
| --- | --- | --- | --- | --- | --- | --- | --- | --- | --- | --- | --- | --- |
|  | *B* | | *SE* | | 95% CIs | *p* | | *B* | *SE* | 95% CIs | | *p* |
| *0.45 threshold* |  |  | |  | |  |  | |  |  |  | |
| SN-CEN FC | 0.23 | 0.08 | | 0.08:0.38 | | 0.004** | 0.03 | | 0.08 | -0.14:0.19 | 0.752 | |
| SN-DMN FC | 0.19 | 0.08 | | 0.04:0.35 | | 0.017* | 0.06 | | 0.09 | -0.11:0.23 | 0.457 | |
| CEN-DMN FC | 0.16 | 0.08 | | -0.00:0.31 | | 0.05* | 0.07 | | 0.08 | -0.10:0.24 | 0.407 | |
| *0.4 threshold* |  |  | |  | |  |  | |  |  |  | |
| SN-CEN FC | 0.22 | 0.08 | | 0.07:0.38 | | 0.006** | 0.04 | | 0.08 | -0.13:0.20 | 0.647 | |
| SN-DMN FC | 0.19 | 0.08 | | 0.03:0.35 | | 0.021* | 0.02 | | 0.09 | -0.15:0.19 | 0.831 | |
| CEN-DMN FC | 0.17 | 0.08 | | 0.01:0.33 | | 0.033* | 0.06 | | 0.09 | -0.11:0.23 | 0.454 | |
| *0.35 threshold* |  |  | |  | |  |  | |  |  |  | |
| SN-CEN FC | 0.25 | 0.08 | | 0.08:0.41 | | 0.004** | 0.01 | | 0.09 | -0.17:0.19 | 0.938 | |
| SN-DMN FC | 0.17 | 0.09 | | -0.01:0.35 | | 0.057^+^ | 0 | | 0.09 | -0.18:0.18 | 0.981 | |
| CEN-DMN FC | 0.21 | 0.09 | | 0.04:0.38 | | 0.016* | 0.04 | | 0.09 | -0.15:0.22 | 0.695 | |
| *0.3 threshold* |  |  | |  | |  |  | |  |  |  | |
| SN-CEN FC | 0.22 | 0.09 | | 0.04:0.40 | | 0.015* | -0.02 | | 0.09 | -0.21:0.17 | 0.849 | |
| SN-DMN FC | 0.16 | 0.09 | | -0.03:0.34 | | 0.094 | 0.01 | | 0.09 | -0.18:0.19 | 0.947 | |
| CEN-DMN FC | 0.19 | 0.09 | | 0.01:0.37 | | 0.042* | -0.03 | | 0.1 | -0.22:0.16 | 0.776 | |
| *0.25 threshold* |  |  | |  | |  |  | |  |  |  | |
| SN-CEN FC | 0.21 | 0.1 | | 0.02:0.40 | | 0.035* | 0.02 | | 0.1 | -0.18:0.22 | 0.834 | |
| SN-DMN FC | 0.19 | 0.1 | | -0.01:0.39 | | 0.067^+^ | -0.03 | | 0.11 | -0.24:0.18 | 0.768 | |
| CEN-DMN FC | 0.14 | 0.1 | | -0.06:0.34 | | 0.175 | -0.05 | | 0.11 | -0.26:0.16 | 0.633 | |
| *0.2 threshold* |  |  | |  | |  |  | |  |  |  | |
| SN-CEN FC | 0.25 | 0.1 | | 0.04:0.46 | | 0.018* | 0.11 | | 0.1 | -0.10:0.31 | 0.308 | |
| SN-DMN FC | 0.24 | 0.11 | | 0.01:0.46 | | 0.043* | 0.27 | | 0.11 | 0.05:0.49 | 0.015 | |
| CEN-DMN FC | 0.09 | 0.11 | | -0.14:0.31 | | 0.441 | -0.02 | | 0.11 | -0.23:0.20 | 0.89 | |
| *0.15 threshold* |  |  | |  | |  |  | |  |  |  | |
| SN-CEN FC | 0.39 | 0.14 | | 0.11:0.67 | | 0.008** | 0.07 | | 0.15 | -0.23:0.38 | 0.631 | |
| SN-DMN FC | 0.19 | 0.17 | | -0.15:0.53 | | 0.276 | 0.41 | | 0.15 | 0.11:0.72 | 0.01* | |
| CEN-DMN FC | 0.14 | 0.16 | | -0.18:0.46 | | 0.383 | -0.1 | | 0.16 | -0.42:0.22 | 0.517 | |

*Note.* Standardised beta coefficients are reported with Standard Errors (SE) and 95% Confidence Intervals (CIs), controlling for age, age^2^, gender, and in-scanner motion. Salience Network (SN), Central Executive Network (CEN), and Default Mode Network (DMN). ^+^*p* < 0.68, **p* < 0.05, ***p* < 0.01

**Table S20**

**Triple Network Connectivity and Neurodevelopmental Difficulties in Comparison Children across Motion Thresholds**

|  | **Hyperactivity/Impulsivity** | | | | | | | **Inattention** | | | | |
| --- | --- | --- | --- | --- | --- | --- | --- | --- | --- | --- | --- | --- |
|  | *B* | | *SE* | | 95% CIs | *p* | | *B* | *SE* | 95% CIs | | *p* |
| *0.45 threshold* |  |  | |  | |  |  | |  |  |  | |
| SN-CEN FC | 0.04 | 0.14 | | -0.25:0.33 | | 0.785 | -0.02 | | 0.14 | -0.30:0.26 | 0.9 | |
| SN-DMN FC | 0.07 | 0.16 | | -0.25:0.38 | | 0.669 | 0.06 | | 0.15 | -0.24:0.36 | 0.698 | |
| CEN-DMN FC | -0.54 | 0.12 | | -0.79:-0.29 | | 6.73e-05*** | -0.12 | | 0.14 | -0.40:0.17 | 0.413 | |
| *0.4 threshold* |  |  | |  | |  |  | |  |  |  | |
| SN-CEN FC | 0.04 | 0.14 | | -0.25:0.33 | | 0.785 | -0.02 | | 0.14 | -0.30:0.26 | 0.9 | |
| SN-DMN FC | 0.07 | 0.16 | | -0.25:0.38 | | 0.669 | 0.06 | | 0.15 | -0.24:0.36 | 0.698 | |
| CEN-DMN FC | -0.54 | 0.12 | | -0.79:-0.29 | | 6.73e-05*** | -0.12 | | 0.14 | -0.40:0.17 | 0.413 | |
| *0.35 threshold* |  |  | |  | |  |  | |  |  |  | |
| SN-CEN FC | -0.05 | 0.15 | | -0.36:0.25 | | 0.721 | 0.01 | | 0.15 | -0.29:0.32 | 0.942 | |
| SN-DMN FC | 0.03 | 0.16 | | -0.28:0.35 | | 0.83 | 0.1 | | 0.15 | -0.21:0.41 | 0.535 | |
| CEN-DMN FC | -0.46 | 0.12 | | -0.71:-0.22 | | 4.05e-4*** | -0.17 | | 0.14 | -0.45:0.10 | 0.21 | |
| *0.3 threshold* |  |  | |  | |  |  | |  |  |  | |
| SN-CEN FC | -0.05 | 0.15 | | -0.36:0.26 | | 0.735 | 0.02 | | 0.15 | -0.28:0.33 | 0.892 | |
| SN-DMN FC | 0.08 | 0.16 | | -0.25:0.40 | | 0.637 | 0.16 | | 0.16 | -0.16:0.48 | 0.318 | |
| CEN-DMN FC | -0.37 | 0.14 | | -0.65:-0.09 | | 0.01** | -0.13 | | 0.15 | -0.43:0.16 | 0.368 | |
| *0.25 threshold* |  |  | |  | |  |  | |  |  |  | |
| SN-CEN FC | -0.05 | 0.16 | | -0.37:0.26 | | 0.726 | 0 | | 0.16 | -0.32:0.33 | 0.991 | |
| SN-DMN FC | 0.08 | 0.17 | | -0.26:0.43 | | 0.635 | 0.12 | | 0.16 | -0.21:0.45 | 0.456 | |
| CEN-DMN FC | -0.42 | 0.14 | | -0.70:-0.13 | | 0.005** | -0.13 | | 0.15 | -0.43:0.16 | 0.365 | |
| *0.2 threshold* |  |  | |  | |  |  | |  |  |  | |
| SN-CEN FC | -0.22 | 0.17 | | -0.55:0.12 | | 0.198 | -0.03 | | 0.16 | -0.37:0.30 | 0.84 | |
| SN-DMN FC | 0.08 | 0.18 | | -0.29:0.44 | | 0.671 | 0.18 | | 0.17 | -0.16:0.53 | 0.294 | |
| CEN-DMN FC | -0.42 | 0.15 | | -0.73:-0.11 | | 0.01** | -0.23 | | 0.16 | -0.56:0.10 | 0.164 | |
| *0.15 threshold* |  |  | |  | |  |  | |  |  |  | |
| SN-CEN FC | 0.16 | 0.27 | | -0.40:0.72 | | 0.552 | -0.11 | | 0.2 | -0.54:0.31 | 0.585 | |
| SN-DMN FC | 0.14 | 0.27 | | -0.43:0.71 | | 0.61 | 0.27 | | 0.23 | -0.22:0.76 | 0.261 | |
| CEN-DMN FC | -0.02 | 0.28 | | -0.61:0.58 | | 0.95 | -0.35 | | 0.23 | -0.83:0.14 | 0.15 | |

*Note.* Standardised beta coefficients are reported with Standard Errors (SE) and 95% Confidence Intervals (CIs), controlling for age, age^2^, gender, and in-scanner motion. Salience Network (SN), Central Executive Network (CEN), and Default Mode Network (DMN). **p* < 0.05, ***p* < 0.01, ****p* < 0.001

**Table S21**

**Group Interaction on CEN-DMN Connectivity and Neurodevelopmental Difficulties across Motion Thresholds**

|  | **Hyperactivity/Impulsivity** | | | | | |
| --- | --- | --- | --- | --- | --- | --- |
| Threshold | *B* | | *SE* | | 95% CIs | *p* |
| 0.45 | 0.65 | 0.16 | | 0.34:0.96 | | 4.31e-05*** |
| 0.4 | 0.66 | 0.16 | | 0.35:0.97 | | 4.34e-05*** |
| 0.35 | 0.64 | 0.17 | | 0.30:0.97 | | 2.09e-4*** |
| 0.3 | 0.5 | 0.19 | | 0.13:0.87 | | 0.008** |
| 0.25 | 0.63 | 0.19 | | 0.26:1.00 | | 0.001** |
| 0.2 | 0.64 | 0.21 | | 0.23:1.05 | | 0.002** |
| 0.15 | 0.06 | 0.4 | | -0.74:0.86 | | 0.882 |

*Note.* The threshold indicates the maximum average framewise displacement*.* Standardised beta coefficients are reported with Standard Errors (SE) and 95% Confidence Intervals (CIs), controlling for age, age^2^, gender, and in-scanner motion. ***p* < 0.01, ****p* < 0.001

**Table S22**

**Adult Network Connectivity and Neurodevelopmental Difficulties in At-Risk Children across Motion Thresholds**

|  | **Hyperactivity/Impulsivity** | | | | | | | **Inattention** | | | | |
| --- | --- | --- | --- | --- | --- | --- | --- | --- | --- | --- | --- | --- |
|  | *B* | | *SE* | | 95% CIs | *p* | | *B* | *SE* | 95% CIs | | *p* |
| *0.45 threshold* |  |  | |  | |  |  | |  |  |  | |
| Ventral SN - CEN | 0.02 | 0.08 | | -0.15:0.18 | | 0.841 | 0.01 | | 0.09 | -0.16:0.19 | 0.883 | |
| Ventral SN - DMN | 0.16 | 0.08 | | 0.00:0.32 | | 0.047* | 0.03 | | 0.08 | -0.13:0.20 | 0.677 | |
| Dorsal SN - CEN | -0.07 | 0.08 | | -0.23:0.09 | | 0.397 | 0.13 | | 0.08 | -0.04:0.30 | 0.125 | |
| Dorsal SN - DMN | 0.13 | 0.08 | | -0.03:0.29 | | 0.111 | 0 | | 0.08 | -0.16:0.16 | 0.992 | |
| CEN - DMN | 0.2 | 0.08 | | 0.05:0.36 | | 0.012* | 0.12 | | 0.08 | -0.04:0.28 | 0.149 | |
| *0.4 threshold* |  |  | |  | |  |  | |  |  |  | |
| Ventral SN - CEN | 0 | 0.08 | | -0.16:0.17 | | 0.981 | 0.01 | | 0.09 | -0.17:0.18 | 0.954 | |
| Ventral SN - DMN | 0.15 | 0.08 | | -0.01:0.32 | | 0.068^+^ | 0.03 | | 0.09 | -0.14:0.20 | 0.755 | |
| Dorsal SN - CEN | -0.07 | 0.08 | | -0.24:0.10 | | 0.402 | 0.1 | | 0.09 | -0.07:0.27 | 0.243 | |
| Dorsal SN - DMN | 0.14 | 0.08 | | -0.03:0.30 | | 0.103 | 0 | | 0.09 | -0.17:0.17 | 1 | |
| CEN - DMN | 0.23 | 0.08 | | 0.08:0.39 | | 0.004** | 0.15 | | 0.08 | -0.02:0.31 | 0.084 | |
| *0.35 threshold* |  |  | |  | |  |  | |  |  |  | |
| Ventral SN - CEN | -0.01 | 0.09 | | -0.19:0.17 | | 0.927 | -0.02 | | 0.1 | -0.21:0.17 | 0.833 | |
| Ventral SN - DMN | 0.12 | 0.09 | | -0.05:0.30 | | 0.165 | 0.01 | | 0.09 | -0.17:0.19 | 0.916 | |
| Dorsal SN - CEN | -0.08 | 0.09 | | -0.26:0.09 | | 0.348 | 0.09 | | 0.09 | -0.10:0.27 | 0.344 | |
| Dorsal SN - DMN | 0.07 | 0.09 | | -0.10:0.25 | | 0.4 | -0.04 | | 0.09 | -0.22:0.15 | 0.694 | |
| CEN - DMN | 0.27 | 0.08 | | 0.10:0.43 | | 0.002** | 0.12 | | 0.09 | -0.06:0.29 | 0.192 | |
| *0.3 threshold* |  |  | |  | |  |  | |  |  |  | |
| Ventral SN - CEN | -0.02 | 0.09 | | -0.20:0.17 | | 0.868 | -0.03 | | 0.1 | -0.22:0.17 | 0.797 | |
| Ventral SN - DMN | 0.11 | 0.09 | | -0.07:0.29 | | 0.247 | -0.05 | | 0.09 | -0.23:0.14 | 0.618 | |
| Dorsal SN - CEN | -0.08 | 0.09 | | -0.26:0.10 | | 0.398 | 0.1 | | 0.1 | -0.10:0.29 | 0.32 | |
| Dorsal SN - DMN | 0.08 | 0.09 | | -0.10:0.27 | | 0.387 | -0.04 | | 0.1 | -0.23:0.15 | 0.705 | |
| CEN - DMN | 0.23 | 0.09 | | 0.06:0.41 | | 0.009** | 0.13 | | 0.09 | -0.06:0.31 | 0.18 | |
| *0.25 threshold* |  |  | |  | |  |  | |  |  |  | |
| Ventral SN - CEN | 0.05 | 0.1 | | -0.16:0.25 | | 0.662 | -0.01 | | 0.11 | -0.23:0.21 | 0.927 | |
| Ventral SN - DMN | -0.02 | 0.1 | | -0.23:0.18 | | 0.813 | -0.04 | | 0.11 | -0.25:0.18 | 0.736 | |
| Dorsal SN - CEN | -0.09 | 0.1 | | -0.29:0.10 | | 0.35 | 0.08 | | 0.11 | -0.13:0.30 | 0.451 | |
| Dorsal SN - DMN | 0 | 0.1 | | -0.20:0.21 | | 0.976 | -0.04 | | 0.11 | -0.26:0.17 | 0.689 | |
| CEN - DMN | 0.14 | 0.1 | | -0.06:0.33 | | 0.164 | 0.11 | | 0.1 | -0.09:0.31 | 0.293 | |
| *0.2 threshold* |  |  | |  | |  |  | |  |  |  | |
| Ventral SN - CEN | 0.09 | 0.11 | | -0.13:0.32 | | 0.405 | 0 | | 0.11 | -0.22:0.22 | 0.989 | |
| Ventral SN - DMN | -0.02 | 0.11 | | -0.25:0.20 | | 0.841 | 0.14 | | 0.11 | -0.08:0.36 | 0.209 | |
| Dorsal SN - CEN | -0.01 | 0.11 | | -0.24:0.21 | | 0.905 | -0.04 | | 0.11 | -0.25:0.17 | 0.712 | |
| Dorsal SN - DMN | -0.03 | 0.11 | | -0.25:0.19 | | 0.807 | -0.01 | | 0.11 | -0.22:0.20 | 0.928 | |
| CEN - DMN | 0.1 | 0.11 | | -0.11:0.32 | | 0.342 | 0.15 | | 0.1 | -0.06:0.36 | 0.151 | |
| *0.15 threshold* |  |  | |  | |  |  | |  |  |  | |
| Ventral SN - CEN | 0.23 | 0.15 | | -0.07:0.54 | | 0.131 | -0.04 | | 0.15 | -0.35:0.27 | 0.791 | |
| Ventral SN - DMN | -0.03 | 0.17 | | -0.37:0.31 | | 0.869 | 0.31 | | 0.16 | -0.01:0.63 | 0.058 | |
| Dorsal SN - CEN | -0.03 | 0.16 | | -0.36:0.29 | | 0.841 | -0.15 | | 0.16 | -0.46:0.17 | 0.349 | |
| Dorsal SN - DMN | -0.14 | 0.17 | | -0.49:0.21 | | 0.428 | 0.19 | | 0.16 | -0.14:0.52 | 0.244 | |
| CEN - DMN | 0.1 | 0.16 | | -0.22:0.42 | | 0.541 | 0.1 | | 0.16 | -0.23:0.42 | 0.556 | |

*Note.* Standardised beta coefficients are reported with Standard Errors (SE) and 95% Confidence Intervals (CIs), controlling for age, age^2^, gender, and in-scanner motion. Salience Network (SN), Central Executive Network (CEN), and Default Mode Network (DMN). ^+^*p* < 0.069, **p* < 0.05, ***p* < 0.01

**Table S23**

**Adult Network Connectivity and Neurodevelopmental Difficulties Comparison Children across Motion Thresholds**

|  | **Hyperactivity/Impulsivity** | | | | | | | **Inattention** | | | |
| --- | --- | --- | --- | --- | --- | --- | --- | --- | --- | --- | --- |
|  | *B* | | *SE* | | 95% CIs | *p* | | *B* | *SE* | 95% CIs | *p* |
| *0.45 threshold* |  |  | |  | |  |  | |  |  |  |
| Ventral SN - CEN | 0.15 | 0.15 | | -0.15:0.46 | | 0.317 | 0 | | 0.16 | -0.31:0.31 | 0.998 |
| Ventral SN - DMN | -0.16 | 0.14 | | -0.45:0.13 | | 0.281 | -0.13 | | 0.14 | -0.41:0.16 | 0.385 |
| Dorsal SN - CEN | 0.14 | 0.15 | | -0.15:0.44 | | 0.334 | 0.07 | | 0.14 | -0.22:0.36 | 0.63 |
| Dorsal SN - DMN | -0.09 | 0.14 | | -0.39:0.20 | | 0.515 | -0.09 | | 0.14 | -0.38:0.19 | 0.506 |
| CEN - DMN | -0.22 | 0.14 | | -0.50:0.05 | | 0.11 | -0.18 | | 0.14 | -0.45:0.10 | 0.196 |
| *0.4 threshold* |  |  | |  | |  |  | |  |  |  |
| Ventral SN - CEN | 0.15 | 0.15 | | -0.15:0.46 | | 0.317 | 0 | | 0.16 | -0.31:0.31 | 0.998 |
| Ventral SN - DMN | -0.16 | 0.14 | | -0.45:0.13 | | 0.281 | -0.13 | | 0.14 | -0.41:0.16 | 0.385 |
| Dorsal SN - CEN | 0.14 | 0.15 | | -0.15:0.44 | | 0.334 | 0.07 | | 0.14 | -0.22:0.36 | 0.63 |
| Dorsal SN - DMN | -0.09 | 0.14 | | -0.39:0.20 | | 0.515 | -0.09 | | 0.14 | -0.38:0.19 | 0.506 |
| CEN - DMN | -0.22 | 0.14 | | -0.50:0.05 | | 0.11 | -0.18 | | 0.14 | -0.45:0.10 | 0.196 |
| *0.35 threshold* |  |  | |  | |  |  | |  |  |  |
| Ventral SN - CEN | 0.11 | 0.15 | | -0.20:0.41 | | 0.475 | 0.05 | | 0.16 | -0.27:0.37 | 0.752 |
| Ventral SN - DMN | -0.15 | 0.15 | | -0.44:0.14 | | 0.307 | -0.14 | | 0.14 | -0.43:0.14 | 0.317 |
| Dorsal SN - CEN | 0.06 | 0.15 | | -0.25:0.36 | | 0.706 | 0.11 | | 0.15 | -0.19:0.40 | 0.464 |
| Dorsal SN - DMN | -0.03 | 0.14 | | -0.32:0.26 | | 0.826 | -0.12 | | 0.14 | -0.40:0.16 | 0.4 |
| CEN - DMN | -0.17 | 0.13 | | -0.44:0.10 | | 0.213 | -0.2 | | 0.13 | -0.47:0.07 | 0.144 |
| *0.3 threshold* |  |  | |  | |  |  | |  |  |  |
| Ventral SN - CEN | 0.07 | 0.15 | | -0.24:0.38 | | 0.661 | 0.03 | | 0.16 | -0.28:0.35 | 0.83 |
| Ventral SN - DMN | -0.15 | 0.15 | | -0.46:0.15 | | 0.311 | -0.14 | | 0.15 | -0.43:0.16 | 0.346 |
| Dorsal SN - CEN | 0.06 | 0.15 | | -0.23:0.35 | | 0.678 | 0.11 | | 0.14 | -0.18:0.40 | 0.454 |
| Dorsal SN - DMN | -0.02 | 0.15 | | -0.32:0.28 | | 0.903 | -0.1 | | 0.14 | -0.39:0.18 | 0.471 |
| CEN - DMN | -0.16 | 0.14 | | -0.45:0.13 | | 0.27 | -0.2 | | 0.14 | -0.48:0.09 | 0.17 |
| *0.25 threshold* |  |  | |  | |  |  | |  |  |  |
| Ventral SN - CEN | -0.02 | 0.16 | | -0.35:0.31 | | 0.907 | -0.02 | | 0.17 | -0.35:0.32 | 0.919 |
| Ventral SN - DMN | -0.11 | 0.16 | | -0.44:0.22 | | 0.496 | -0.14 | | 0.15 | -0.45:0.17 | 0.375 |
| Dorsal SN - CEN | 0.01 | 0.15 | | -0.30:0.32 | | 0.951 | 0.09 | | 0.15 | -0.22:0.39 | 0.574 |
| Dorsal SN - DMN | -0.02 | 0.16 | | -0.34:0.31 | | 0.916 | -0.12 | | 0.15 | -0.43:0.18 | 0.418 |
| CEN - DMN | -0.2 | 0.16 | | -0.52:0.12 | | 0.205 | -0.23 | | 0.15 | -0.53:0.08 | 0.143 |
| *0.2 threshold* |  |  | |  | |  |  | |  |  |  |
| Ventral SN - CEN | 0.08 | 0.17 | | -0.27:0.43 | | 0.643 | -0.02 | | 0.18 | -0.38:0.33 | 0.889 |
| Ventral SN - DMN | -0.11 | 0.18 | | -0.47:0.24 | | 0.525 | -0.14 | | 0.16 | -0.48:0.19 | 0.39 |
| Dorsal SN - CEN | 0.07 | 0.17 | | -0.28:0.42 | | 0.706 | 0.03 | | 0.17 | -0.32:0.37 | 0.871 |
| Dorsal SN - DMN | -0.07 | 0.17 | | -0.41:0.28 | | 0.705 | -0.12 | | 0.16 | -0.45:0.21 | 0.457 |
| CEN - DMN | -0.24 | 0.17 | | -0.58:0.10 | | 0.164 | -0.33 | | 0.17 | -0.66:0.01 | 0.058^+^ |
| *0.15 threshold* |  |  | |  | |  |  | |  |  |  |
| Ventral SN - CEN | 0 | 0.28 | | -0.59:0.59 | | 0.992 | -0.15 | | 0.24 | -0.65:0.35 | 0.54 |
| Ventral SN - DMN | 0.16 | 0.25 | | -0.35:0.68 | | 0.514 | -0.1 | | 0.21 | -0.54:0.35 | 0.648 |
| Dorsal SN - CEN | 0.08 | 0.29 | | -0.54:0.69 | | 0.799 | -0.33 | | 0.21 | -0.76:0.11 | 0.133 |
| Dorsal SN - DMN | 0.17 | 0.26 | | -0.37:0.70 | | 0.522 | -0.01 | | 0.21 | -0.45:0.42 | 0.953 |
| CEN - DMN | 0.12 | 0.28 | | -0.47:0.71 | | 0.664 | -0.48 | | 0.18 | -0.87:-0.10 | 0.016* |

*Note.* Standardised beta coefficients are reported with Standard Errors (SE) and 95% Confidence Intervals (CIs), controlling for age, age^2^, gender, and in-scanner motion. Salience Network (SN), Central Executive Network (CEN), and Default Mode Network (DMN). ^+^*p* < 0.059, **p* < 0.05

**Table S24**

**Group Interaction on Adult CEN-DMN Connectivity and Neurodevelopmental Difficulties across Motion Thresholds**

|  | **Hyperactivity/Impulsivity** | | | | | |
| --- | --- | --- | --- | --- | --- | --- |
| Threshold | *B* | | *SE* | | 95% CIs | *p* |
| 0.45 | 0.37 | 0.16 | | 0.06:0.68 | | 0.021* |
| 0.4 | 0.39 | 0.16 | | 0.07:0.70 | | 0.016* |
| 0.35 | 0.37 | 0.17 | | 0.03:0.70 | | 0.032* |
| 0.3 | 0.35 | 0.18 | | -0.00:0.69 | | 0.051^+^ |
| 0.25 | 0.4 | 0.19 | | 0.03:0.77 | | 0.036* |
| 0.2 | 0.42 | 0.21 | | 0.01:0.83 | | 0.046* |
| 0.15 | -0.31 | 0.4 | | -1.11:0.49 | | 0.444 |

*Note.* The threshold indicates the maximum average framewise displacement*.* Standardised beta coefficients are reported with Standard Errors (SE) and 95% Confidence Intervals (CIs), controlling for age, age^2^, gender, and in-scanner motion. ^+^*p* < 0.052, **p* < 0.05

**Table S25**

**Network Connectivity Differences between the Samples over Motion Thresholds**

|  | At-Risk | Comparison | t-test | | | Regression | | | |
| --- | --- | --- | --- | --- | --- | --- | --- | --- | --- |
|  | *M* (*SD*) | *M* (*SD*) | *t* | *D* | *p* | *β* | *SE* | CI | *p* |
| *0.45 threshold* |  |  |  |  |  |  |  |  |  |
| SN - CEN | 0.11 (0.17) | 0.06 (0.16) | -2.13 | -0.33 | 0.034* | -0.2 | 0.15 | -0.51:0.10 | 0.194 |
| SN - DMN | 0.17 (0.20) | 0.19 (0.21) | 0.74 | 0.11 | 0.46 | 0.08 | 0.16 | -0.23:0.40 | 0.598 |
| CEN - DMN | 0.19 (0.15) | 0.17 (0.16) | -0.82 | -0.12 | 0.411 | -0.03 | 0.16 | -0.34:0.28 | 0.858 |
| *0.4 threshold* |  |  |  |  |  |  |  |  |  |
| SN - CEN | 0.11 (0.17) | 0.06 (0.16) | -1.94 | -0.3 | 0.054^+^ | -0.19 | 0.15 | -0.49:0.12 | 0.229 |
| SN - DMN | 0.16 (0.19) | 0.19 (0.21) | 0.84 | 0.13 | 0.404 | 0.09 | 0.16 | -0.23:0.40 | 0.595 |
| CEN - DMN | 0.19 (0.15) | 0.17 (0.16) | -0.72 | -0.11 | 0.473 | -0.03 | 0.16 | -0.34:0.28 | 0.835 |
| *0.35 threshold* |  |  |  |  |  |  |  |  |  |
| SN - CEN | 0.09 (0.16) | 0.05 (0.15) | -1.77 | -0.28 | 0.079 | -0.23 | 0.16 | -0.55:0.10 | 0.17 |
| SN - DMN | 0.15 (0.18) | 0.18 (0.21) | 0.99 | 0.15 | 0.325 | 0.06 | 0.17 | -0.27:0.40 | 0.702 |
| CEN - DMN | 0.18 (0.15) | 0.17 (0.17) | -0.08 | -0.01 | 0.94 | 0.05 | 0.17 | -0.27:0.38 | 0.755 |
| *0.3 threshold* |  |  |  |  |  |  |  |  |  |
| SN - CEN | 0.10 (0.15) | 0.05 (0.16) | -2.02 | -0.32 | 0.045* | -0.29 | 0.17 | -0.62:0.04 | 0.086 |
| SN - DMN | 0.14 (0.18) | 0.20 (0.20) | 1.77 | 0.28 | 0.079 | 0.17 | 0.17 | -0.16:0.51 | 0.311 |
| CEN - DMN | 0.17 (0.14) | 0.16 (0.15) | -0.38 | -0.06 | 0.704 | -0.03 | 0.17 | -0.37:0.31 | 0.872 |
| *0.25 threshold* |  |  |  |  |  |  |  |  |  |
| SN - CEN | 0.10 (0.15) | 0.02 (0.13) | -3.31 | -0.59 | 0.001*** | -0.55 | 0.18 | -0.90:-0.20 | 0.002** |
| SN - DMN | 0.15 (0.18) | 0.19 (0.22) | 1.22 | 0.2 | 0.223 | 0.11 | 0.18 | -0.24:0.47 | 0.529 |
| CEN - DMN | 0.18 (0.14) | 0.15 (0.16) | -0.99 | -0.16 | 0.324 | -0.1 | 0.18 | -0.46:0.26 | 0.58 |
| *0.2 threshold* |  |  |  |  |  |  |  |  |  |
| SN - CEN | 0.11 (0.15) | 0.02 (0.16) | -3.15 | -0.57 | 0.002** | -0.49 | 0.18 | -0.85:-0.13 | 0.008** |
| SN - DMN | 0.17 (0.16) | 0.19 (0.22) | 0.67 | 0.12 | 0.503 | 0.04 | 0.19 | -0.34:0.42 | 0.823 |
| CEN - DMN | 0.17 (0.13) | 0.15 (0.15) | -0.96 | -0.17 | 0.338 | -0.1 | 0.19 | -0.48:0.28 | 0.602 |
| *0.15 threshold* |  |  |  |  |  |  |  |  |  |
| SN - CEN | 0.09 (0.15) | 0.01 (0.13) | -2.58 | -0.62 | 0.012* | -0.59 | 0.23 | -1.05:-0.13 | 0.013* |
| SN - DMN | 0.18 (0.17) | 0.22 (0.20) | 0.89 | 0.21 | 0.374 | 0.12 | 0.24 | -0.37:0.61 | 0.62 |
| CEN - DMN | 0.17 (0.13) | 0.10 (0.12) | -2.05 | -0.51 | 0.044 | -0.43 | 0.25 | -0.92:0.06 | 0.086 |

*Note.* The threshold indicates the maximum average framewise displacement*.* Standardised beta coefficients are reported with Standard Errors (SE) and 95% Confidence Intervals (CIs), controlling for age, age^2^, gender, and in-scanner motion. Cohen’s D (*D*), Salience Network (SN), Central Executive Network (CEN), Default Mode Network (DMN). ^+^*p* < 0.055, **p* < 0.05, ***p* < 0.01, ****p* < 0.01

**Table S26**

**Network Connectivity Differences in ADHD over Motion Thresholds**

|  | ADHD | Comparison | t-test | | | Regression | | | |
| --- | --- | --- | --- | --- | --- | --- | --- | --- | --- |
|  | *M* (*SD*) | *M* (*SD*) | *t* | *D* | *p* | *β* | *SE* | CI | *p* |
| *0.45 threshold* |  |  |  |  |  |  |  |  |  |
| SN - CEN | 0.16 (0.13) | 0.06 (0.16) | -3.08 | -0.7 | 0.003** | -0.56 | 0.24 | -1.04:-0.08 | 0.023* |
| SN - DMN | 0.23 (0.17) | 0.19 (0.21) | -0.78 | -0.18 | 0.44 | -0.15 | 0.26 | -0.67:0.36 | 0.554 |
| CEN - DMN | 0.24 (0.08) | 0.18 (0.16) | -1.73 | -0.45 | 0.087 | -0.18 | 0.27 | -0.71:0.36 | 0.512 |
| *0.4 threshold* |  |  |  |  |  |  |  |  |  |
| SN - CEN | 0.16 (0.13) | 0.06 (0.16) | -3.08 | -0.7 | 0.003** | -0.56 | 0.24 | -1.04:-0.08 | 0.023* |
| SN - DMN | 0.23 (0.17) | 0.19 (0.21) | -0.78 | -0.18 | 0.44 | -0.15 | 0.26 | -0.67:0.36 | 0.554 |
| CEN - DMN | 0.24 (0.08) | 0.18 (0.16) | -1.73 | -0.45 | 0.087 | -0.18 | 0.27 | -0.71:0.36 | 0.512 |
| *0.35 threshold* |  |  |  |  |  |  |  |  |  |
| SN - CEN | 0.17 (0.11) | 0.05 (0.15) | -3.47 | -0.88 | 0.001*** | -0.75 | 0.25 | -1.26:-0.25 | 0.004** |
| SN - DMN | 0.22 (0.17) | 0.19 (0.21) | -0.72 | -0.17 | 0.476 | -0.24 | 0.27 | -0.78:0.31 | 0.39 |
| CEN - DMN | 0.23 (0.08) | 0.17 (0.17) | -1.51 | -0.41 | 0.135 | -0.13 | 0.27 | -0.68:0.41 | 0.629 |
| *0.3 threshold* |  |  |  |  |  |  |  |  |  |
| SN - CEN | 0.15 (0.13) | 0.05 (0.16) | -2.77 | -0.69 | 0.007** | -0.64 | 0.26 | -1.16:-0.12 | 0.017* |
| SN - DMN | 0.21 (0.17) | 0.21 (0.19) | 0.03 | 0.01 | 0.98 | -0.15 | 0.28 | -0.70:0.40 | 0.592 |
| CEN - DMN | 0.22 (0.08) | 0.16 (0.15) | -1.75 | -0.5 | 0.085 | -0.21 | 0.29 | -0.78:0.37 | 0.472 |
| *0.25 threshold* |  |  |  |  |  |  |  |  |  |
| SN - CEN | 0.20 (0.11) | 0.02 (0.13) | -5.75 | -1.55 | 2.52E-07*** | -1.31 | 0.25 | -1.80:-0.82 | 1.52E-06*** |
| SN - DMN | 0.21 (0.17) | 0.21 (0.20) | -0.17 | -0.04 | 0.863 | -0.16 | 0.29 | -0.74:0.43 | 0.597 |
| CEN - DMN | 0.19 (0.10) | 0.15 (0.16) | -1.05 | -0.3 | 0.299 | -0.05 | 0.3 | -0.66:0.55 | 0.857 |
| *0.2 threshold* |  |  |  |  |  |  |  |  |  |
| SN - CEN | 0.20 (0.10) | 0.03 (0.16) | -4.24 | -1.3 | 7.47E-05*** | -1.13 | 0.28 | -1.70:-0.56 | 2.00E-4*** |
| SN - DMN | 0.21 (0.18) | 0.20 (0.22) | -0.32 | -0.09 | 0.747 | -0.22 | 0.32 | -0.86:0.41 | 0.489 |
| CEN - DMN | 0.21 (0.08) | 0.14 (0.16) | -1.72 | -0.55 | 0.091 | -0.32 | 0.33 | -0.97:0.33 | 0.334 |
| *0.15 threshold* |  |  |  |  |  |  |  |  |  |
| SN - CEN | 0.24 (0.06) | 0.01 (0.15) | -3.87 | -1.96 | 4.86E-04*** | -1.52 | 0.43 | -2.40:-0.63 | 0.002** |
| SN - DMN | 0.20 (0.19) | 0.25 (0.18) | 0.74 | 0.28 | 0.466 | 0.25 | 0.48 | -0.74:1.23 | 0.61 |
| CEN - DMN | 0.24 (0.09) | 0.09 (0.13) | -2.94 | -1.25 | 0.006** | -0.91 | 0.44 | -1.82:-0.00 | 0.049* |

*Note.* The threshold indicates the maximum average framewise displacement*.* Standardised beta coefficients are reported with Standard Errors (SE) and 95% Confidence Intervals (CIs), controlling for age, age^2^, gender, and in-scanner motion. Cohen’s D (*D*), Salience Network (SN), Central Executive Network (CEN), Default Mode Network (DMN). **p* < 0.05, ***p* < 0.01, ****p* < 0.01

**Exploratory Analyses**

We conducted post-hoc tests to explore the group interaction of hyperactivity/impulsivity on CEN-DMN connectivity. We first investigated the hypothesis that the functional segregation of the CEN and DMN may diverge with age in the at-risk sample relative to the comparison sample. We assessed whether CEN-DMN connectivity was significantly associated with age or age^2^, whilst controlling for gender, in-scanner motion, and age^2^ or age respectively.

In the adult networks, we found that CEN-DMN connectivity seemed to linearly increase with age in the comparison sample (*n* = 53, *β* = 0.3, *p* = 0.057) rather than quadratically change (*β* = 0.00, *p* = 0.979). Whereas in the at-risk sample, CEN-DMN connectivity did not linearly increase with age (*n* = 169, *β* = -0.12, *p* = 0.142), but seemed to quadratically decrease in younger children before flattening in older children (*β* = 0.16, *p* = 0.051). These neurodevelopmental differences were displayed using Locally Weighted Scatterplot Smoothing (LOWESS) in statsmodels 0.12.1 (see Figure S1). LOWESS is a nonparametric fitting technique that can estimate nonlinear relationships by calculating weighted averages between neighbouring data points.

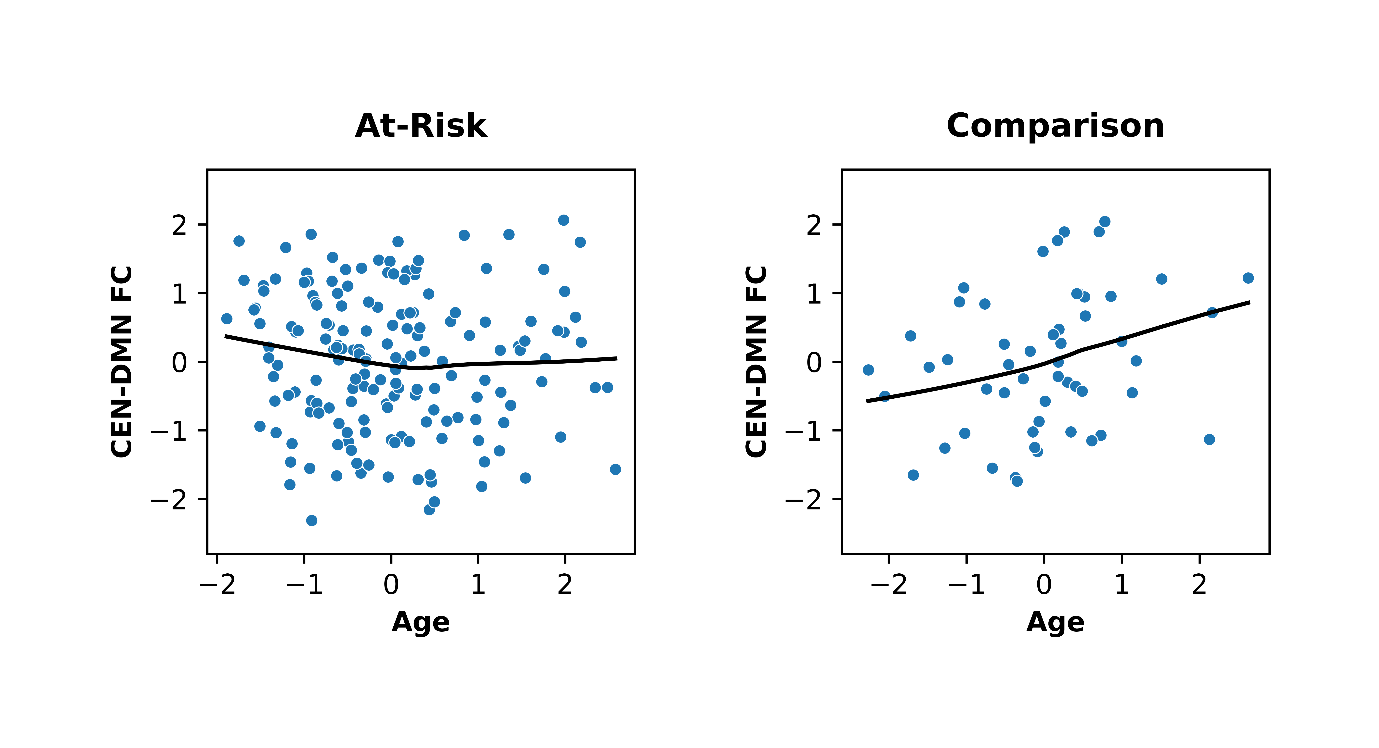

Figure S1. LOWESS nonparametric association between age and CEN-DMN functional connectivity between the two samples, after gender and in-scanner motion have been regressed out.

Recently, in a large sample of children recruited from the Adolescent Brain and Cognitive Development study, greater CEN-DMN connectivity was shown to predict greater cognitive ability in some children and worse in others (Ellwood-Lowe, Whitfield-Gabrieli, & Bunge, 2021). Therefore, we investigated whether increasing CEN-DMN connectivity could be cognitively advantageous in our sample of comparison children. We used age and gender standardised scores from the Matrix Reasoning subtest of the Wechsler Abbreviated Scale of Intelligence II (Wechsler, 2011). We then ran two linear regressions with matrix reasoning as the independent variable and CEN-DMN connectivity and hyperactivity/impulsivity as the dependent variables, while controlling for age, age^2^, and gender, as well as in-scanner motion for the regression on CEN-DMN connectivity.

In comparison children, matrix reasoning significantly positively associated with CEN-DMN connectivity (*n* = 53, *r* = 0.3, *p* = 0.027, *β* = 0.27, *p* = 0.051) and negatively associated with hyperactivity/impulsivity (*r* = -0.35, *p* = 0.011, *β* = -0.35, *p* = 0.012). Whereas in the at-risk sample, matrix reasoning was not associated with CEN-DMN connectivity (*n* = 166, *r* = -0.07, *p* = 0.353, *β* = -0.07, *p* = 0.355) or hyperactivity/impulsivity (*r* = -0.01, *p* = 0.900, *β* = -0.02, *p* = 0.705). There was a significant group*reasoning interaction effect on CEN-DMN connectivity, demonstrating that matrix reasoning was only positively associated with CEN-DMN connectivity in the comparison sample (*n* = 219, *β* = -0.2, *p* = 0.013).

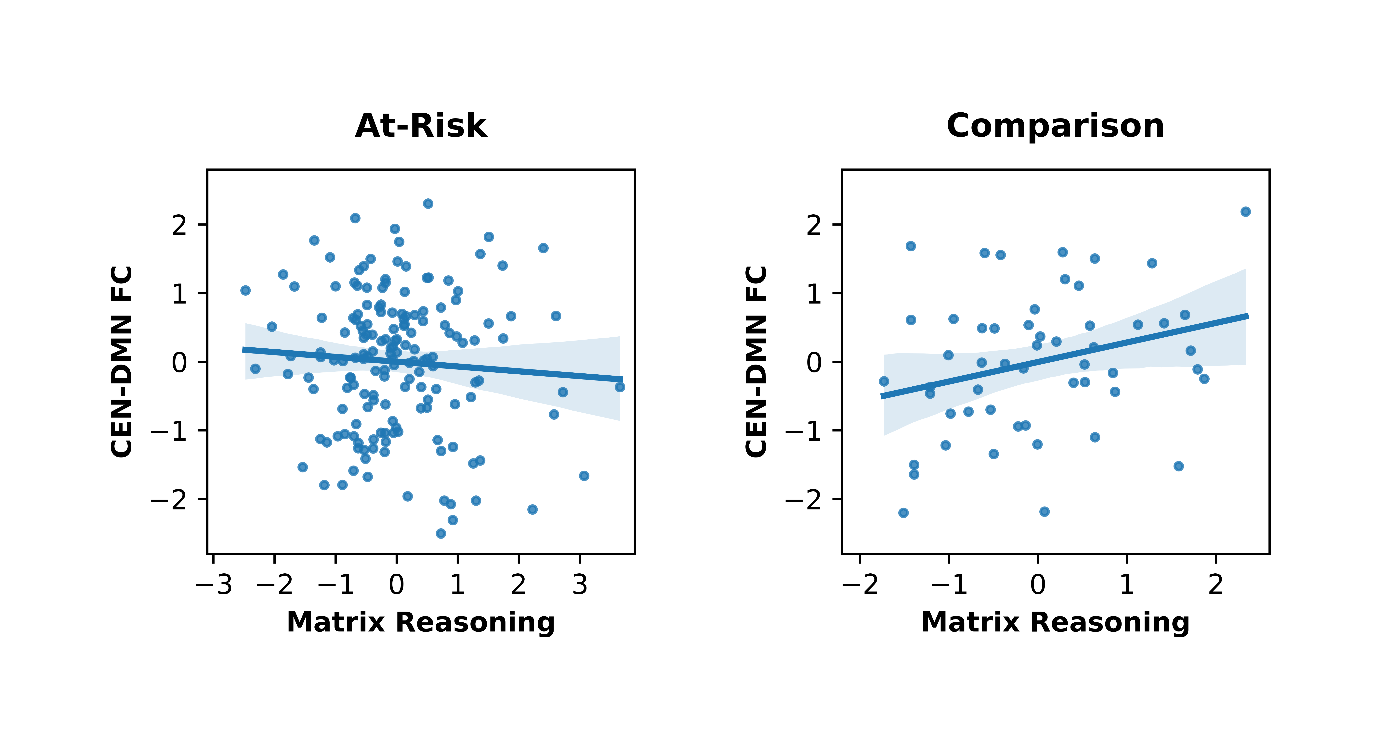

Figure S2. Association between Matrix Reasoning and CEN-DMN functional connectivity between the two samples, after age, age^2^, gender, and in-scanner motion have been regressed out.
